# The Societal Value of Vaccines: Expert-Based Conceptual Framework and Methods Using COVID-19 Vaccines as A Case Study

**DOI:** 10.1101/2022.12.02.22283046

**Authors:** Manuela Di Fusco, Diana Mendes, Lotte Steuten, David E Bloom, Michael Drummond, Katharina Hauck, Jonathan Pearson-Stuttard, Rachel Power, David Salisbury, Adrian Towse, Julie Roiz, Gabor Szabo, Jingyan Yang, Kinga Marczell

## Abstract

Health technology assessments (HTAs) of vaccines typically focus on the direct health benefits to individuals and healthcare systems. COVID-19 highlighted the widespread societal impact of infectious diseases and the value of vaccines in averting adverse clinical consequences and in maintaining or resuming social and economic activities. Using COVID-19 as a case study, this research work aimed to set forth a conceptual framework capturing the broader value elements of vaccines and to identify appropriate methods to quantify value elements not routinely considered in HTAs. A two-step approach was adopted combining a targeted literature review and three rounds of expert elicitation based on a modified Delphi method, leading to a conceptual framework of 30 value elements related to broader health effects, societal and economic impact, public finances, and uncertainty value. When applying the framework to COVID-19 vaccines in post-pandemic settings, 13 value elements were consensually rated highly important by the experts for consideration in HTAs. The experts reviewed over 10 methods that could be leveraged to quantify broader value elements and provided technical forward-looking recommendations. Limitations of the framework and the identified methods were discussed. This study supplements on-going efforts aimed towards a broader recognition of the full societal value of vaccines.

## 1. Introduction

The dynamic changes in the social landscape brought by the coronavirus disease 2019 (COVID-19) have shown the multiplicity of harms and broad impacts that infectious diseases may bring to society. The public health and societal impact of the COVID-19 pandemic and the associated economic and social fallouts have been far-reaching [1-5]. The measures taken by governments to mitigate the pandemic, such as mass vaccination, social-distancing, mask wearing, and lockdowns have also directly or indirectly affected individuals and their social interactions, health systems and the world economy [6-9].

Traditional health technology assessment (HTA) focuses on a narrow list of health-related effects, including reduction in mortality and morbidity, lowering healthcare costs and resource use, and in certain HTA jurisdictions, increasing productivity of patients and their caregivers [10-12].

A growing body of evidence advocates that vaccines have benefits and externalities that extend beyond these traditional categories, hence they may be systematically undervalued in HTAs [13-29]. Over the past decade, several generic and vaccine-specific value frameworks have been developed using a broad societal perspective [13-29]. More recently, researchers concurred that the COVID-19 pandemic showed the limits of current HTAs and contributed to broadening the view of what constitutes vaccines value in HTA [13, 17, 22, 24, 28, 30]. Bell *et al*., [13] identified gaps between the broader value elements included in early vaccination-specific value frameworks and the value elements generally being considered in HTAs of vaccines, which resulted in a synthesised framework called the ‘Broader Value of Vaccines’ (BRAVE) Framework.

While the broader value of vaccines is being increasingly recognised and theoretically understood, there is limited research and consensus on the methods to use to measure broader value elements and integrate them meaningfully in economic evaluations and decision making [26, 29].

Building on the existing body of evidence, expert inputs, and the growing literature on the effects of COVID-19 vaccination, the aims of this study were threefold: (1) to create a consensus-based conceptual framework capturing the broader value of vaccination, (2) to gather expert recommendations and views on challenges related to analytical approaches measuring these benefits, (3) to assess the applicability of the framework and the identified methods for evaluations of COVID-19 vaccines, with a focus on post-pandemic settings.

## 2. Materials and Methods

The study adopted a two-step approach combining a targeted literature review (TLR) and an expert elicitation exercise.

### Targeted Literature Review (TLR)

The objectives of the TLR were: (1) to identify general and vaccine-specific value frameworks characterising vaccine value, (2) to describe the economic and societal outcomes impacted by COVID-19, and (3) to identify methods to include such impacts in HTA processes.

Considering the breadth of the research topics underpinning these objectives and the vastness of the COVID-19 literature, an all-encompassing systematic review would have covered almost the entirety of the vaccine literature. We therefore opted to leverage the latest research by Brassel *et al*., [17] and Bell *et al*., [13] to understand the current state of vaccine value assessments, and to conduct targeted searches to identify relevant articles on the impact of COVID-19 and the effects of COVID-19 vaccination. The targeted searches were conducted between 14 December, 2021 and 7 April, 2022 in PubMed and Google Scholar. The TLR integrated two main types of evidence: observational studies and models, with the latter being especially helpful to identify relevant analytical approaches.

The latest vaccine frameworks and the evidence on the economic and societal outcomes impacted by COVID-19 were analysed and synthesised in an ‘Impact Inventory’. A gap analysis was conducted to identify elements of the impact inventory that are not currently recognised in HTAs, with a focus on the UK and the US. Finally, available methodologies for their quantification were identified and summarised. An appraisal of the literature was conducted before the expert elicitation phase. The appraisal considered both the quality of the evidence linking the value elements to COVID-19 and/or vaccines, and the ability to incorporate the value elements in economic evaluations, expressed as the feasibility to monetise them (Appendix C, Table S2).

### Expert Elicitation

The purpose of the expert elicitation phase was fourfold: (1) to review and validate the TLR findings, (2) to leverage the ‘Impact Inventory’ to develop a consensus-based conceptual framework of broad vaccine value elements, (3) to seek consensus on the broader value elements of the framework to be prioritised in post-pandemic COVID-19 vaccines assessments, and (4) to gather recommendations and challenges on methods quantifying those prioritised value elements.

Similar to Bell *et al*., [13] the expert elicitation phase was based on a modified Delphi, a well-established technique combining existing evidence with multiple expert perspectives to identify gaps and priorities for research and decision making [31, 32].

The expert elicitation was performed in three rounds (Figure 1), and leveraged a questionnaire developed based on the TLR findings (Appendix A). Round 1 was an individual expert elicitation. Round 2 and Round 3 expert elicitations were conducted during a virtual panel discussion (Panel 1). Panel 1 was followed by another virtual panel (Panel 2) which was a qualitative discussion to review the methods identified in the TLR. While the two panel discussions allowed group interaction, all the elicitations were individual and anonymous.

**Figure 1.**
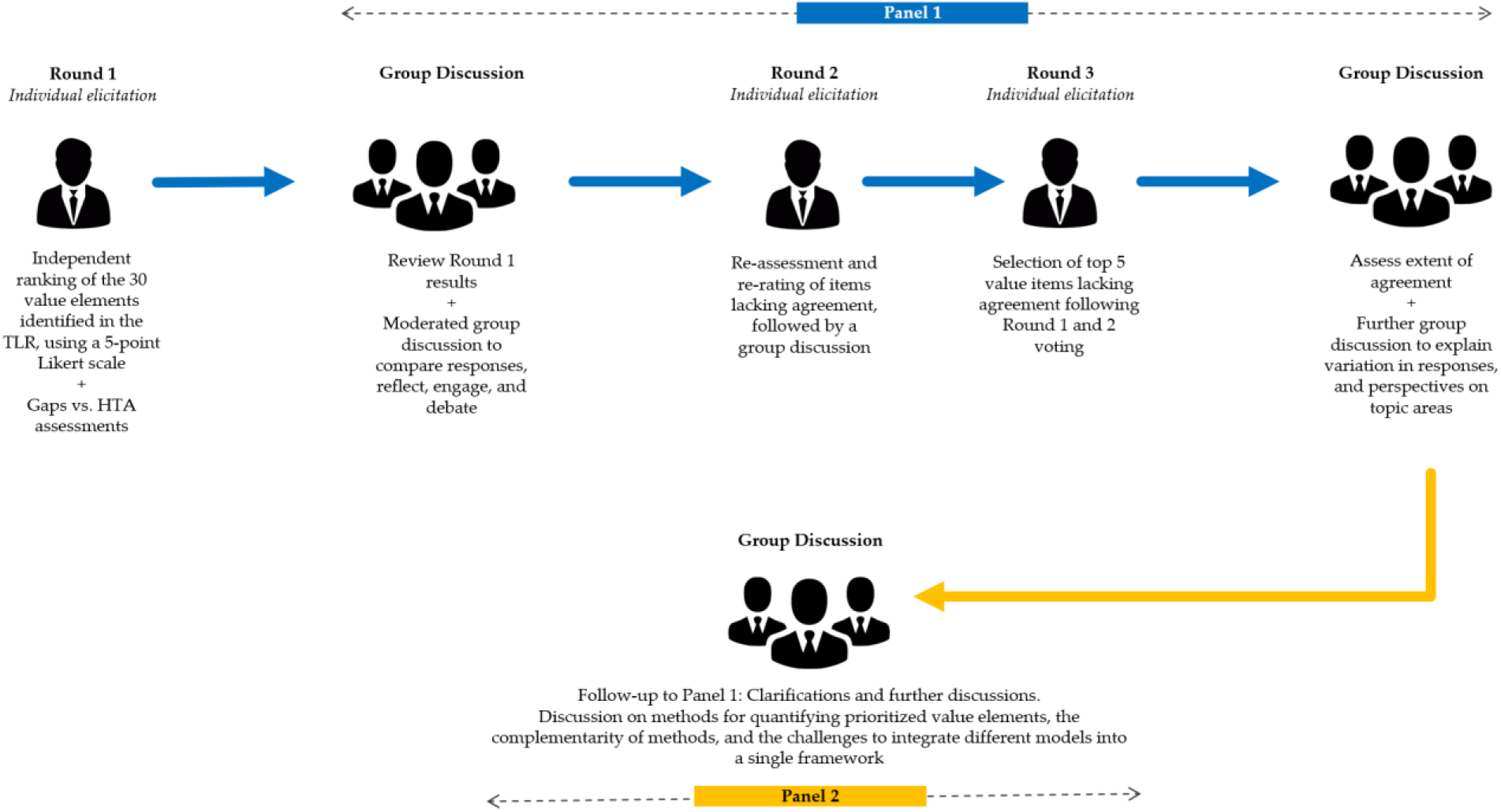
Expert Elicitation using a modified Delphi method. HTA, health technology assessment; TLR, targeted literature review

Given the complexity and breadth of the research topics, the panel eligibility criteria included in-depth knowledge of HTA methods and decision making as well as expertise in applicable areas such as health economics, public health, immunisation, outcomes research, infectious disease epidemiology, HTA and health policy. A total of nine experts with comprehensive and authoritative knowledge were engaged in the discussions and the individual expert elicitations. The panel included an expert in direct elicitation of patients and provider benefit-risk trade-off preferences, and a patient advisory group representative. A distinguished expert in health economics acted as the moderator for both panels who also contributed to the discussions while participating in the individual expert elicitation rounds as a facilitator.

The panel sample size was defined based on the BRAVE example [13], prior research and guidelines [31-36], and considerations of response rate, group dynamics, and stability of responses. A 70% level of agreement was considered appropriate in Delphi guidelines and prior Delphi studies with similar panel size [31-39]. Hence, our study aimed for this threshold with slight modifications in either direction as the number of responders changed slightly across rounds, as further detailed in the Results section. Round 1: Individual Expert Elicitation

In Round 1, each expert was provided with a summary of the TLR findings, including the ‘Impact Inventory’, and was invited to take part in a survey via email (Appendix A). The experts were asked to rate the value elements that, conceptually, were priority for inclusion in the economic evaluations of COVID-19 vaccines, as well as the quality of evidence underpinning their inclusion. They also commented on the overall feasibility of their inclusion or the rationale for their exclusion in existing HTA frameworks. The questionnaire used a 5-point Likert scale along with a few open-ended questions that asked the experts to state their view independently of the perspective recommended by the HTA bodies (Appendix A).

The experts then were asked if the methods identified in the TLR were suitable for inclusion of broader value elements within economic evaluations of COVID-19 vaccines. Finally, they were asked about other potential methods and relevant examples and best practices.

Round 1 was followed by two half-day virtual panel discussions: Panel 1 in May 2022, and Panel 2 in June 2022.

### Panel 1 (Rounds 2 and 3)

Panel 1 focused on building an expert-informed conceptual framework encompassing broader vaccine value elements, and on gathering consensus on the value elements to be prioritised for inclusion in economic evaluations of COVID-19 vaccines, with a focus on post-pandemic settings.

During Panel 1, two main sets of results were presented to the experts: (1) the TLR findings, including the ‘Impact Inventory’ and a gap analysis versus existing HTA frameworks, and (2) the Round 1 survey results, analysed prior to the virtual discussion. The presentation of these results was followed by a moderated group discussion to reflect on Round 1 responses, debate interpretations and uncover differences in opinions. Following clarifications and discussion, the experts were invited to take part in Round Via live online polls, Round 2 repeated the prioritisation process with focus on the value elements for which consensus was not achieved in Round 1. A subsequent group discussion was conducted to discuss the new value elements that were prioritised. In Round 3, the experts were asked to select the top five value elements for which consensus was not reached during the prior rounds based on their relevance or priority for inclusion in economic evaluations. The experts collectively reviewed the resulting consensus, as well as a summary of the main topics and expert considerations that emerged from Panel 1.

### Panel 2

Panel 2 was a focused semi-structured technical discussion on the methods and challenges to capture and quantify the broader value elements prioritised during Panel 1. Panel 2 aimed to discuss challenges and recommendations for methodological approaches enabling the integration of the broader effects of vaccination in economic evaluations, with a focus on COVID-19 vaccines. There was no formal voting or prioritisation. The experts were asked to review existing analytical approaches, and to share examples and solutions to overcome some of the challenges associated with these approaches. The results of the discussion were analysed qualitatively based upon a content analysis of notes and transcripts. A descriptive summary was organised around each value element.

## 3. Results

### 3.1. Findings of the Targeted Literature Review

The TLR resulted in the identification of more than 15 value frameworks, with eight of them specific for vaccines, indicating significant progress and strong advocacy to advance vaccine value assessment using a broad societal perspective [13, 15-25, 27-29].

The TLR identified a relatively long list of value elements that are not captured in traditional vaccine value assessments [10-12]. A total of 30 identified value elements were described in an ‘Impact Inventory’ (Appendix B), further detailed in the next section.

The results of a gap analysis versus existing frameworks showed that several of the value elements were presented in different levels of detail and classification in various existing frameworks, as shown in Figure 2 and Figure S1. One of the latest vaccine-specific value frameworks, BRAVE [13], captured all the key categories of value attributed to vaccines in prior frameworks. As such, recognising the overlaps among the frameworks, BRAVE was used as the reference inventory of broader value elements to build upon and potentially augment.

**Figure 2.**
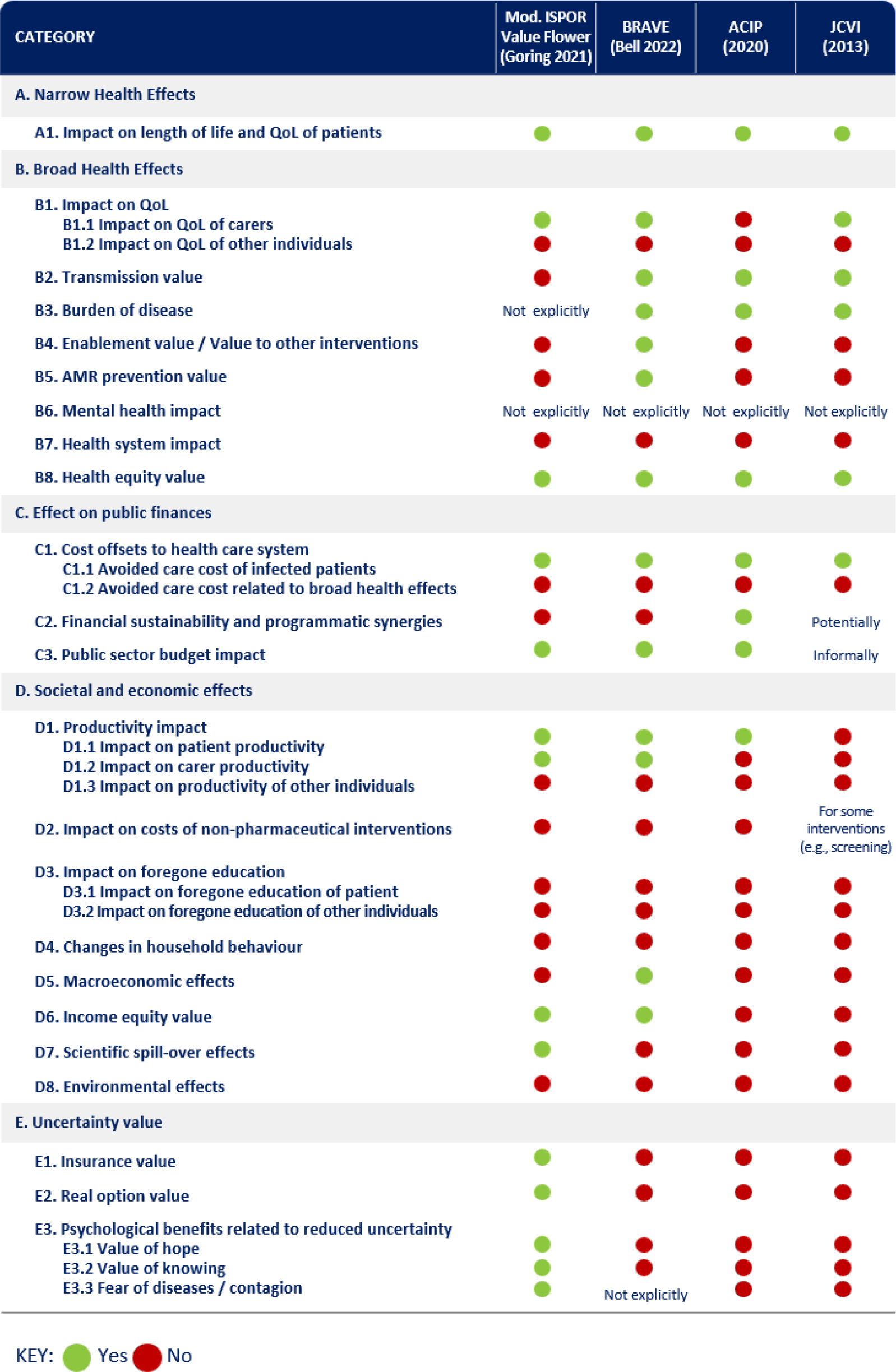
Inclusion of Value Elements in Key Value Frameworks. ACIP, Advisory Committee on Immunization Practices (US); BRAVE, Broader Value of Vaccines; JCVI, Joint Committee on Vaccination and Immunisation (UK); QOL, Quality of Life Note: Vaccine Anxiety was added to the framework ex-post based on the panel discussions.

#### 3.1.1. Identified Value Elements

The ‘Impact Inventory’ (Appendix B) lists the identified value elements. To preserve comprehensiveness, the first four categories of effects from BRAVE were retained. These followed an order from the narrow perspective towards a broader one, and from health-related outcomes towards economic and societal outcomes: narrow (A) and broad (B) health effects were followed by the cost and income outcomes for the healthcare system (C1 and C2), the public sector (C3), and the society (D). The narrow category (A) included value elements related to the impact of vaccination on the health of the vaccinated subjects. The broad health effects category (B) included effects related to the impact of vaccination on the health of the unvaccinated subjects and other relevant individuals such as caregivers. Category C was concerned with the impact of vaccination on the health system. Category D focused on the broad societal and economic impact of vaccination, beyond the healthcare system, and spanning to broad macroeconomic effects. Category E was added based upon the TLR results and especially building on the novel value elements of the ‘ISPOR Value Flower’ [24]. This category of effects included those on the uncertainty of health and economic outcomes, including the psychological impact of vaccination (i.e., value of hope, value of knowing, fear of diseases/contagion), and insurance value.

One of the initial findings from the TLR was that most of the quantifiable value elements and outcomes impacted by COVID-19 were captured in BRAVE. Several elements that were considered relevant to COVID-19 but not explicitly listed in BRAVE were added to the framework, including mental health impact, impact on health system, effect on public finances, impact on direct costs of non-pharmaceutical interventions (NPIs), impact on foregone education, changes in individual and household behaviour, income equity value, scientific spill-over effects, environmental effects, and uncertainty value. Some of these elements might be implicitly included in BRAVE (e.g., broad health outcomes may include mental health impact), but they were separately mentioned because of their importance and/or prominent consideration in the COVID-19 research. Most of these elements have been primarily associated with the pandemic and the resulting NPIs, as opposed to the COVID-19 cases. However, it was also considered that these elements may prove to be of relevance for future immune escape COVID-19 variants, or for other diseases that are associated with large-scale containment measures.

#### 3.1.2. Identified Quantification Methods

The literature review subsequently identified methods to quantify the impact of COVID-19 or vaccination on the value elements that are usually not being considered in economic evaluations of vaccines. The amount and strength of the identified evidence and the perceived ability of incorporating the elements in quantitative evaluations varied across value elements. Appendix C and D summarise these analyses and considerations, whereas Appendix E describes all such identified quantification methods in detail, along with the experts’ views collected during the panel discussions.

Narrow health effects (A) were not in the scope of the TLR, as the impacts on length of life and quality of life (QoL) of patients are usually included in traditional cost-effectiveness analysis of vaccines, as such, several well-known methods exist for the quantification of these effects. The broader health effects (B) including the mental health impact of the COVID-19 pandemic and related social restrictions, the overload on health systems, the disproportional direct/indirect health burden of disadvantaged groups in the society associated with the pandemic, were well documented [39-46]. However, limited evidence was identified to assess the monetary value of vaccination through its impact on mental health or considering the health equity aspect of the pandemic or vaccination [47-49]. While certain elements of the public finance impact of the pandemic (C) have been estimated for both the US and UK, no study was identified calculating the full public finance impact or the return on investment of the COVID-19 vaccines [18, 50, 51].

For the societal and economic effects (D), literature assessing the various aspects of the pandemic’s macroeconomic impact was abundant, mainly concentrating on GDP and employment as outcome measures [40, 42, 52-61]. Contrarily, limited published evidence was available estimating the impact of vaccination on the direct cost of NPIs [30]. Finally, for uncertainty value (E), increasing COVID-19 vaccination rates have shown psychological benefits, measured by lower levels of anxiety, worry, displeasure, and depression in the US [62]. However, no identified study had attached monetary value to the psychological value of vaccination.

### 3.2. Results of the Expert Elicitations

#### 3.2.1. Panel Characteristics

Panel 1 was composed of a total of nine experts, of whom eight (89%) responded to the individual elicitation Round 1. The experts had worked in healthcare sector or research for 29 years on an average (min-max: 12-47); most worked in academic or governmental institutions; with seven doing research primarily in the UK setting, four in the US, and four on an international scale. A total of seven experts participated in Round 2, and a total of nine experts participated in Round 3.

Based on these panel compositions, consensus in Round 1 was defined as >75% (six out of eight) of participant panellists providing either high rating (4 or 5) or a low rating (1 to 2) to a value element. In Round 2, consensus was defined similarly, using ≥ 67% (six out of nine) as threshold.

Given the focus of Panel 2 on the methods to capture broad value, the composition of Panel 2 was narrowed down to the seven experts with health economics expertise that attended Panel 1.

#### 3.2.2. Conceptual Value Framework and Considerations

The experts provided several important conceptual considerations before and after the expert elicitations rounds.

Among the initial remarks, the experts pointed out that the profound impacts of COVID-19 and COVID-19 vaccines showed the limits of the existing HTAs for vaccines in capturing their full public benefits. Despite increasing recognition of the broader value of vaccination, narrow direct benefits remain the focus of vaccine HTAs, with potential implications on public investments and immunisation policies. The experts pointed out that the staggering effects of COVID-19 and the broad benefits associated to COVID-19 vaccines provide an opportunity to advance perspectives on the importance of accounting for the full benefits of vaccines in HTAs.

The experts mentioned that, during identification and collation of elements reflecting the full value of a vaccine, ideally, a framework should integrate elements that are conceptually appropriate for any vaccine evaluation. The relevance and magnitude of impact of each value element of the framework would then differ on a case-by-case basis, according to the vaccine and settings under assessment. In this regard, the experts agreed that value elements are strongly dependent on the specific vaccine and circumstances it is being evaluated under, specifically considering pandemic versus endemic settings, short versus long analytic time horizons, direct and indirect effects. For example, the direct costs of NPIs were considered primarily related to the pandemic nature of COVID-19 and less of a concern for endemic phases. Value elements strongly related to NPIs, such as lockdowns, are expected to be highly relevant and have a substantial impact in an evaluation of primary vaccinations in a pandemic setting, in the context of low levels of population-based immunity, rising infections, limited vaccine and treatment options, and voluntary and non-voluntary contact restrictions. In an endemic setting, the benefits of vaccination should be balanced with additional factors defining the counterfactual scenario against which the impacts on value elements are expected to be quantified, such as the existence of effective and affordable therapies and the presence of vaccine comparators. However, the pandemic versus endemic differentiation was not considered binary: as COVID-19 transitions to an endemic disease, some of the indirect impacts experienced during the pandemic phase, such as large productivity impacts associated with quarantines, or disruptions in production, or foregone education associated to school closures are still present, have lingering or spill-over effects. The improvements in health brought by the COVID-19 vaccines can reap long-term benefits, including strengthening economic stability, influencing individual and household behaviours such as fertility, and improving educational outcomes. These benefits can be relevant to be considered for future evaluation of boosters and potentially new formulations against new variants, even if with a lower quantitative impact than previously. As such, there was unanimous consensus in recommending that assessments of COVID-19 vaccines in either pandemic or endemic settings should be conducted using a societal perspective.

For the purpose of developing a comprehensive framework of all value elements that, in principle, could co-exist and be considered in vaccine evaluations, the panel agreed to first investigate the conceptual importance of value items that should be part of it. The assessment of the ability to measure and incorporate them in an economic evaluation should be a second step. It was emphasised that just focusing upfront on aspects that are easy to measure and for which good quality evidence is available could lead to neglect of important value elements. In cases where a value element is considered important from a societal perspective, the causal impact of vaccination on it is conceptually established, and the expected value of information is high, stakeholders and the scientific community should be encouraged to refine the methodological toolset and collect data to fill in the evidence gaps. Filling evidence gaps to capture broad value in vaccine assessments requires resources, but the evidence generated may align incentives for the industry and research, and may improve the quality of decision making.

The experts considered it pragmatic to build and expand on BRAVE. While reviewing the list of value elements included in the ‘Impact Inventory’, expanding on BRAVE, the experts pointed out that the elements listed represent mutually dependent concepts. Depending on how they are labelled and described, they can be sometimes interpreted as overlapping. For instance, psychological benefits of vaccination may potentially and partially translate to impact on mental health as measured by incidence of depression and anxiety; productivity impact influences public sector budget through transfers and taxes, and both are related to macroeconomic outcomes; impact on patient QoL is part of the burden of disease. The experts highlighted that, while a comprehensive framework inevitably includes overlapping value elements, assessments of specific vaccines in specific contexts should be designed in a way that excludes overlaps and double counting. It is theoretically possible to limit double counting by presenting evidence on overlapping items separately rather than summing them quantitatively into a single value metric. However, this is arguably not a preferred approach in an HTA context as it does not lead to an ultimate metric of cost-effectiveness that could be compared against predefined thresholds. The panel recommended that value elements that cannot be quantified and included in the assessment without overlaps should be either considered for exclusion from the analysis or analysed in a qualitative way to support deliberations. However, they emphasised that, in practical applications, overlaps are less likely to arise in assessments based on well-defined integrated epidemiological and economic models, potentially with relatively short analytic time horizons.

The experts pointed out that the assessment of the impact of a disease and the evaluation of a vaccine should balance positive and negative aspects and consider the ‘net’ effect of each value element. In the case of COVID-19, for example, negative externalities of COVID-19 impact on the health system include diversion of resources, leading to delayed diagnoses and treatments for other diseases, and reduced uptake of other immunisations. On the other hand, some of the positive externalities which could arguably be linked to both COVID-19 vaccination and NPIs include air quality improvement during lockdowns and reduced infection rates of other respiratory diseases such as respiratory syncytial virus infection or influenza [63-66]. Similarly, it was emphasised that the potential negative effects of vaccination should be considered and spelled out separately in the framework. These include disutility and work productivity loss related to adverse events, vaccine anxiety leading to fear and uncertainty over long-term effects of vaccination, as well as potential negative effects derived from compensatory behavioural adjustments post-vaccination, for example related to increased social interactions based on the perception of being protected from infection. While attaching a quantitative value to psychological effects of vaccination was considered difficult, conceptually these aspects were considered to have an impact on uptake, infection numbers and potentially quality of life. Moreover, they were considered especially relevant by the panel in mandatory vaccination programs, where some people may have to take the vaccine despite having a negative subjective valuation for it. As a result of these considerations, vaccine anxiety was added under the category E3 of the ‘Impact Inventory’, and the following revisions were made: mortality and QoL impact of potential adverse events related to vaccination were captured more explicitly in category A1; attitude towards risk of infection and infection control measures, and willingness to vaccinate against other diseases were added under the D4 category of behavioural changes, mental health and foregone education were captured more explicitly too. The value framework is visualised in Figure 3.

**Figure 3.**
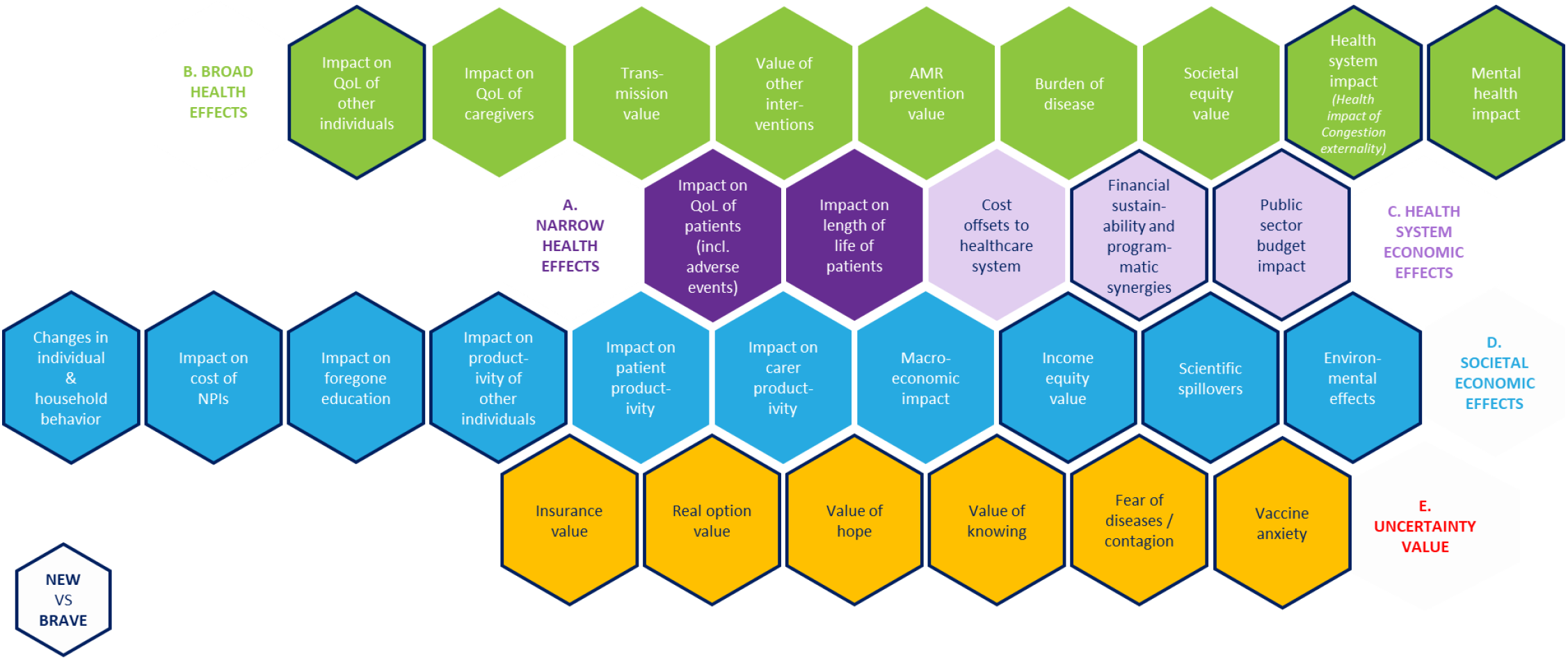
Visualised Vaccine Value Framework.

#### 3.2.3. Prioritisation of Value Elements for COVID-19 Vaccine Evaluations

The voting results related to the prioritisation of value elements for inclusion in COVID-19 vaccine value assessments based on their conceptual appropriateness are presented in the ‘Priority for Inclusion’ columns in Figure 4. The value elements are listed in the order of their priority for inclusion in assessment, wherein, consensually rated elements were highlighted in green, and elements substantially lacking consensus in yellow.

**Figure 4.**
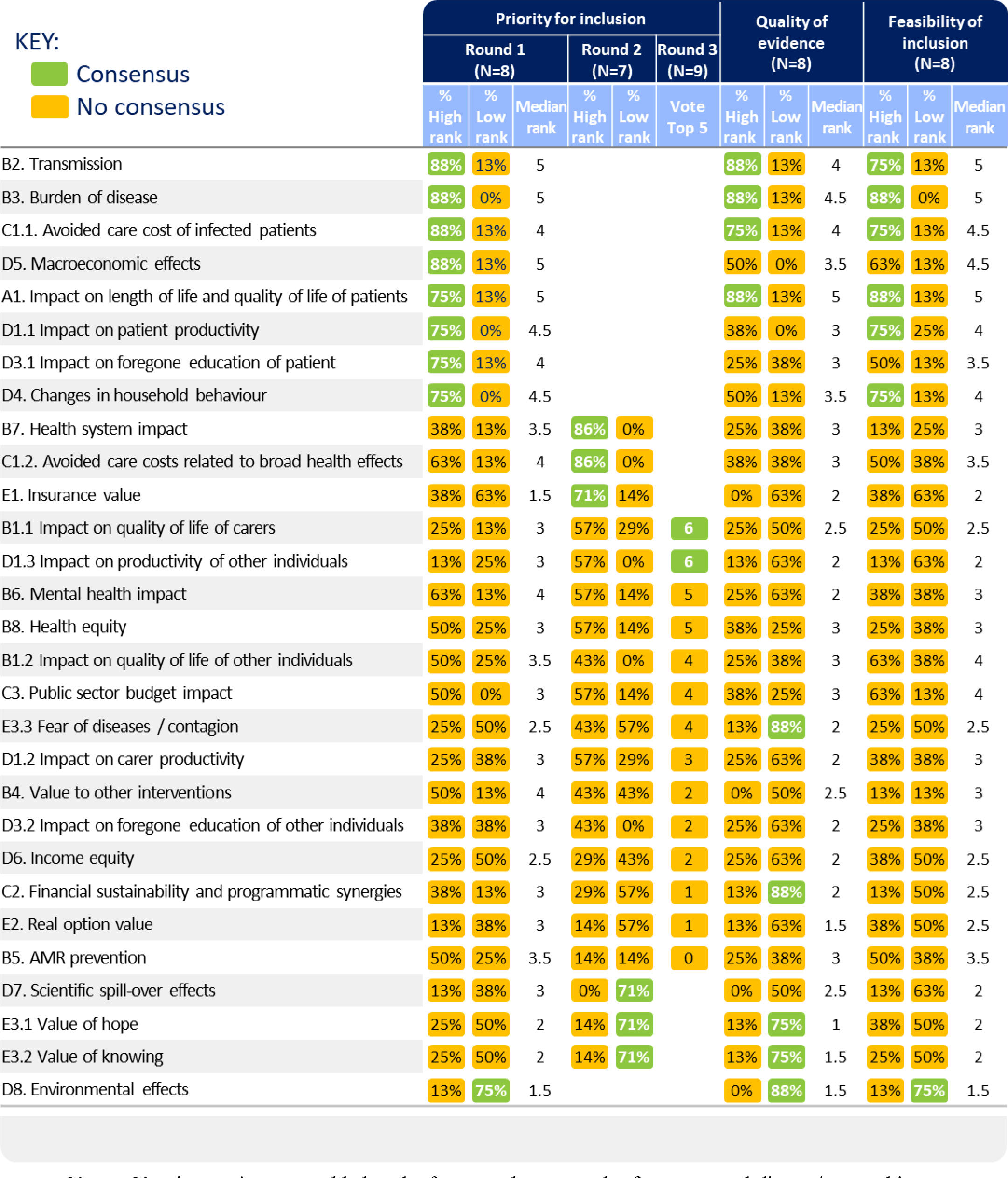
Results of Expert Elicitation Rounds. Note – Vaccine anxiety was added to the framework as a result of expert panel discussions and is not covered in this table.

In Round 1 and 2 combined, 11 value elements were consensually rated as of ‘high importance’ (rated 4 or 5 on the 5-point Likert scale) for inclusion in COVID-19 vaccine assessments, including three elements from the broader health effects (B), four from the societal and economic impact (D), two from public finances (C), insurance value (E), and impact on length of life and quality of life (QoL) of patients (A). The 11 elements were: transmission value, burden of disease, avoided care cost of infected patients, macroeconomic effects, impact on length and QoL of patients, impact on patient productivity, impact on foregone education of patients, changes in individual and household behaviour, health system impact, avoided care costs related to broad health effects, and insurance value.

After Round 2, the experts discussed the criteria considered to prioritise value elements, whether the value element is quantifiable in addition to its likely magnitude, whether feasible to compare to the cost of vaccination rollouts, and consideration of a combination of what matters to people, governments, and what is known about the impact of COVID-19.

The voting results related to subsequent questions on the quality of evidence and the ability/feasibility of inclusion in economic evaluations are presented in the corresponding columns in Figure 4 and Table S3 in Appendix E. Most of the value elements prioritised for inclusion in HTA evaluations by the experts were characterised by high-quality and high-feasibility. Figure 4 shows that, among all the items prioritised for inclusion, the ones that were consensually rated as ‘high’ and also being backed up by ‘high-quality evidence’ and a ‘high feasibility for inclusion’, were the ones that were already included in many or all vaccine value assessments: transmission value, burden of disease, avoided care cost of infected patients, and impact on length and QoL of patients. Insurance value was prioritised in Round 2, although with a relatively low score on evidence and feasibility. High priority and feasibility items without consensus on high evidence included impact on patient productivity, and changes in individual and household behaviour. The experts discussed many aspects related to individual and household behaviour, such as decisions related to fertility, consumption, leisure, savings, living arrangements, attitudes towards risk of infection, willingness to vaccinate against other diseases, impact on quality of life related to isolation at home. Some of these aspects are captured via other value elements in the framework (e.g., impact on savings under macroeconomic effect). In those instances, behavioural changes constitute a channel through which value elements are impacted by COVID-19 and vaccination.

One aspect that was considered especially important was the prevalence elastic demand for prevention, whereby increased vaccination uptake in the population could lead to lower individual risk avoiding behaviour. Increased risk-taking following vaccination may lead to more social contacts and potentially more infections than what would have been observed without the behavioural response.

Value elements that had consistently ‘low’ (rated 1 or 2) scores for priority of inclusion were environmental effects, value of knowing, value of hope, and scientific spill-over effects. For the first three, this assessment was coupled with a consensually low rating for quality of evidence, while environmental effects also got a low rating for the feasibility of inclusion.

Besides the 11 value elements prioritised in Rounds 1 and 2, during Round 3, six out of nine participants rated the ‘impact on QoL of carers’ and ‘impact on productivity of other individuals’ as being one of the top five elements among the ones lacking consensus in Rounds 1 and 2. Two value elements were not consensually prioritised but were emphasised as not receiving sufficient attention in current assessments: impact on caregiver productivity, health equity and related aspects of social and economic equity. From a patient’s perspective, it was highlighted that informal carers may face both productivity and health impacts as a result of their caring responsibilities. A vaccine that reduces the need for informal care can generate economic savings related to unpaid care, as well as may contribute to overall health equity.

During the discussions, it was highlighted that while antibiotic usage has been impacted by COVID-19, the significance and impact of this effect on the development and transmission of AMR was not yet measured. The limited evidence contributed to a relatively low overall rating for ‘AMR prevention’.

### 3.2.4. Evaluation of Identified Quantification Methods

The review of quantification methods focused on selected value elements prioritised for inclusion in COVID-19 vaccine evaluations. The Panel 2 discussion prioritised the review of methods for the following value elements: impact on work productivity, macroeconomic effects, impact on foregone education and health system impacts. The Appendix E presents the results from Round 1 voting on the appropriateness of the identified quantification methods for the elements that were considered relevant by Panel 1.

The list of methods that were identified included well-established approaches for quantifying the impact on work productivity, namely the friction cost and the human capital methods [67]. The experts discussed the pros and cons of both and commented that the human capital approach captures lost income due to mortality/morbidity associated with a disease at an individual level and it is practical for diseases leading to short-term sickness absence, such as acute respiratory diseases. However, some of the aspects of productivity are not generally considered in these types of assessments, such as the fact that losing one’s job and getting re-employed is associated with a substantial loss of firm-specific human capital. Aligned to prior observations related to the need to balance positive and negative effects, the experts pointed out that the productivity loss linked to the administration and management of adverse events of vaccination should be accounted for, too. Finally, the experts also highlighted that, in a pandemic setting, the estimation of the productivity impact needs to consider individuals beyond patients and caregivers. The effect of mandatory quarantine periods, the higher risk of a longer leave and unemployment may be mitigated by recent adjustment to new working arrangements and cost-savings arising from avoided commuting to the office.

The macroeconomic impact emerged as a key topic in the panel discussions and recent literature [13, 26]. The TLR identified a long list of mutually dependent macroeconomic aggregates and indicators. Some of the quantified outcomes included tourism income, production, and GDP in total and at sectoral level, GDP components (e.g., consumption, household income), employment including hours worked – partially overlapping with ‘productivity effect’ as included in HTA, domestic and foreign investment, financial market indicators (e.g., stock market indices), and societal welfare measures. The experts pointed to existing research focusing on GDP estimates [40], and highlighted that, when assessing the macroeconomic impact of vaccines through avoiding NPIs, focusing on the aggregate GDP measure (or gross value added) as the only macroeconomic outcome measure was considered as an appropriate approach for synthesising macroeconomic impact. This single measure avoids the need to look at multiple interrelated macroeconomic outcomes, minimising risk of double counting. However, as GDP is not sensitive to distributional outcomes, impact on health equity and income inequalities through social welfare measures, were also recommended. Building *de novo* macroeconomic or combined epidemiological and macroeconomic models can be helpful from a scenario analysis perspective to assess whether results point in the same direction, but such complex models do not seem appropriate for HTA purposes and for endemic settings, due to their complexity, data needs and inherent uncertainties.

The economic impact of education loss has been assessed in terms of lost schooldays (due to illness and/or school closures), which have been linked to test scores, lost future income and GDP loss. The Organisation for Economic Co-operation and Development (OECD) approach for assessing the impact of lost education on individual income and GDP was considered straightforward and worthy of conduction [56]. However, the experts highlighted that the method is not sensitive to distributional consequences, which were considered important. The impact of lost schooling on the distribution of test scores is relevant both from an equity perspective, and also because the share of high achievers is a key driver in economic growth. The panel suggested an assessment of the impact across different education levels. From a technical perspective, the experts mentioned that such assessments should consider the effects of policy measures aiming to mitigate the impact of school-closures on cognitive outcomes, such as alternative teaching methods (summer camps, online programs). Microsimulation models estimating the impact of lost education on individuals’ future productivity [58] were considered complex from a HTA perspective.

Multiple sources documented the impact of COVID-19 pandemic stretching the resilience of health systems to provide non COVID-19-related care in the same manner as pre-pandemic level. These effects have been measured by simple pre- and post-March 2020 comparisons in the time series with a variety of outcome variables such as length of waiting lists, length of waiting times, number of treatments and screenings performed (e.g., for cancer), hidden needs (an estimate on the number of people that need care but have not yet come forward to receive care), number of incomplete patient pathways, and excess deaths [43, 46]. The experts considered it appropriate to evaluate the healthcare resources by their opportunity cost, as done by Brassel *et al*., [18], as opposed to their accounting cost (e.g., cost of a hospital bed). Brassel *et al*., [18] estimated the opportunity cost of treating a patient instead of another in an excess demand situation. For assessing the monetary value on the health system, the opportunity costs were proxied by the net monetary benefit foregone to treat a vaccine-preventable outcome instead of treating a patient from the waiting list. The experts discussed alternative methods estimating the value of non-COVID-19 life years saved using excess deaths from non-COVID-19 causes and the value of a life year [42], or, alternatively, of a statistical life. They discussed that the excess deaths as a measure for the indirect mortality impact of COVID-19—including but not limited to the health system impact—can be controversial, and it should only be assumed during time periods when demand for health services exceeds capacity. Hence, in forward looking analyses, for instance, a multiplier capturing the relationship between the number of intensive care unit (ICU) cases and excess inpatient deaths could be used for predicting excess deaths based on case numbers. The experts also pointed out that, ideally, the applications of this method should consider interdependencies across care settings and the underlying background risks of existing conditions in the population of interest. For example, missed check-ups or delayed treatments for conditions treated in the primary care sector may result in deterioration and accelerated increase in severity of such conditions that may ultimately require inpatient care. Finally, the experts suggested exploring assessments of QoL decrements, too, especially for deteriorating conditions that are unlikely to cause short-term death and can cause suffering to unattended patients waiting to receive care.

The experts highlighted challenges in measuring the impact of individual and household behaviour. They pointed out that this category may include a long list of poorly documented and quantifiable effects related to ‘household production’. For example, increased time devoted to household childcare responsibilities as a result of school closures.

A broad economic evaluation can capture health, economic, and social impacts. As a direction for future research in this field, the experts commented that, when structuring the economic assessment of a vaccine, an integrated epidemiological, economic and social model, despite being complex for standard HTA, can help conceptualising the framework and evaluating the relationship among value elements. They highlighted that, when presenting evidence on the value of vaccination, it is important to assess both parameter uncertainty and model uncertainty by pursuing alternative modelling approaches and presenting their comparative results transparently. Whenever possible, comparing conclusions from trial-data based estimates against real world evidence is recommended. When considering the inclusion of conceptually important but non-monetary value items in economic evaluations, they suggested to conduct a multi-criteria decision analysis with expert involvement. Finally, conducting comparisons of vaccine value estimates based on evidence-based narrow versus broader perspectives would provide insights into the value of information to be potentially generated for future assessments.

## 4. Discussion

The COVID-19 pandemic and its profound impact on society has reignited the debate about the scope of vaccine value assessments. This research work aimed to contribute to this discussion and advance the growing consensus that vaccines generate value beyond direct effects for the vaccinated individuals and healthcare systems only. Using COVID-19 as a case study and leveraging a mixed-method approach combining a TLR and expert elicitation, this research work introduced a comprehensive list of value elements associated with vaccination for potential consideration either in endemic or pandemic settings. When applying the framework to COVID-19 vaccines in a post-pandemic setting, 13 value elements were consensually rated highly important by the experts for consideration in HTA evaluations. The expert consultation process resulted in an clear consensus on the need to expand existing narrow vaccine HTA frameworks to better emphasise the value of vaccination from a broad societal perspective. The experts discussed the need for a framework that captures value elements reflecting the impact of vaccination on patients, caregivers, and rest of the society. For practical applications, this framework should include value elements that are potentially operationalisable and likely quantifiable, but should not necessarily be confined to items for which good-quality quantitative evidence is available already. The conceptual and quantitative importance of value elements are expected to vary across diseases, country settings (private, public), endemic and pandemic context, and the availability of treatments and alternative vaccine platforms.

The resulting framework (Figure 4) was based on both vaccine and non-vaccine specific frameworks, and showed similarities with previous studies, as several of the value elements prioritised for broader recognition were discussed in expert panels by Bell *et al*., [13], Postma *et al*., [26], and Asukai *et al*., [14] with the latter being focused on COVID-19 therapeutics. High-rated value elements not currently considered in vaccine evaluations but covered in the BRAVE framework included macroeconomic effects, and cost consequences of health system impact. Value elements not included in the BRAVE framework but rated highly by experts were impact on foregone education of patients, health (QoL) consequences of health system impact, insurance value, and changes in individual and household behaviour—specifically, the tendency of people to engage in more social contact as a response to decreased infection risk due to vaccination. Some of these value elements were recommended for prioritisation in Postma *et al*., [26] such as macroeconomic impact, health system externalities, and foregone education, which was covered under productivity impact. Asukai *et al*., [14] discussed equity, disease severity, insurance value, scientific and family spill-over. Environmental impact—or, in Postma *et al*., more specifically, carbon footprint—received low scores in both expert groups. Psychological effects of vaccination and scientific spill-overs were valued less in this study’s expert panels than in the analysis reported by Postma *et al*., [26]. Similarly, while some experts emphasised the importance of considering the equity aspect of vaccination, there was no consensus on its prioritisation in this expert panel, contrary to its high support in Postma *et al*., [26], and in Asukai *et al*., [14] in a pandemic setting. Further, although AMR prevention (B5) was rated generally low, driven by availability and quality of evidence, some of the experts emphasised its quantification feasibility and potential future value, should more evidence emerge.

The framework proposes new value elements that have not been covered previously, such as vaccine anxiety, health, QoL, and economic impact on individuals other than patients and caregivers. The relevance, quality of evidence and possibility of inclusion of the value elements in future vaccine evaluations were assessed by the experts, with a focus on COVID-19 vaccines in a post-pandemic setting. Conceptual considerations of including new value elements in assessments were also discussed, including the importance of generating theoretical and empirical evidence related to value items that are highly relevant but are currently overlooked due to evidence gaps and the lack of appropriate quantification methods.

Differently from prior studies [13, 14], this study collected and reviewed currently available quantification methods to assign value to the identified elements. The strengths and limitations of quantification methods were discussed with the experts both from a general, academic perspective, and in the context of HTA evaluations. The conceptual and methodological challenges related to the strong relationship between value elements and resulting overlaps were analysed, and recommendations were made about potential approaches to limit double counting.

The results of this study should be considered in the context of several limitations.

As emerged during the panel discussions, several aspects of the framework should be interpreted with caution. The framework brings together value elements identified in the TLR and in different existing vaccine and non-vaccine frameworks, where each value element has a different level of detail, validation and tangibility. Seen altogether, the value elements are not mutually exclusive, several of them are interdependent and overlapping, hence they should not be seen simply as additive to one another. Further research should focus on improving their descriptions further and delineating their interdependencies. Quite similarly, many of the methods identified by the TLR were considered relevant and suitable for capturing the impacts of vaccine by the panel, although several challenges exist for their use and there is scope for improving them. In addition, the conceptual and quantitative importance of value elements may vary across vaccines, diseases, settings and time horizons. This framework should be used and re-evaluated on a case-by-case basis, to identify value elements that are applicable to the specific intervention under evaluation. Further, this study focuses on the need for a broad perspective on Vaccine HTAs, however, similar criteria would apply to other health interventions (e.g., pharmaceutical drugs, medical devices) and non-health interventions with health implications, while prioritising the allocation of health budget.

The composition of our panel mainly represented the US and UK and, despite its diversity, it cannot provide full representation of societal preferences. Hence, the results of this research may not be generalisable to countries with different or less established HTA process and should be interpreted in the context of the limitations of the panel composition and size. While the response rates across the three rounds were relatively high (88% in Round 1, 77% in Round 2, 100% in Round 3), the stability of responses might have been affected by the differences in participants and their own interpretations of the topics, especially in the early rounds.

Fundamentally, the size of a Delphi panel can range from 3 to 80 participants [34-36], and our panel selection criteria and size were consistent with similar prior efforts (Bell *et al*., [13] included 10 experts in the panel). While we feel that the group of experts was heterogenous and highly knowledgeable in this research field, we acknowledge that any small-sample qualitative research has limited generalisability. It is possible that a larger panel and/or a different composition could have led to a different final set of recommendations. For example, the results would probably have been different if the panel included a larger number of HTA specialists and a different mix of experiences in the topic of this research work. Further, while the lack of full anonymity between panellists contradicts one of the basic rules in the Delphi method, a lack of discussion could also hamper clarification of disagreements. Hence, our Delphi was modified to include communication among experts and, to minimise the biases from dominance or group pressure, survey responses were always kept anonymous.

Finally, given the large and growing amount of literature, we took a pragmatic approach and focused efforts on a targeted literature review. A substantial amount of evidence was available on the impact of COVID-19 on indirect health outcomes, loss of schooldays, macroeconomic impact, public finances, antibiotic use, and on certain environmental outcomes; however, research evaluating certain value elements and evidence directly relating COVID-19 vaccination to these broad outcomes was scarce and relatively uncertain. As evidence on the impacts of COVID-19 and the COVID-19 vaccines is still accumulating, and methods for quantifying the effects are constantly evolving, further research and expert debate is warranted for this important and complex topic.

## 5. Conclusions

The impact of vaccination extends beyond the direct health effects on vaccinated individuals and healthcare systems. Using COVID-19 as a case study and a mixed-method approach, this research work introduces a conceptual framework of elements to consider when assessing the value of vaccines.

From the exercise emerged an unequivocal consensus on the importance of assessing the value of COVID-19 vaccines using a societal perspective. Several value elements were consensually rated highly important by the experts for consideration in HTA evaluations of COVID-19 vaccines in a post-pandemic setting. Moreover, recommendations and challenges on methods to quantify those were summarised. The findings of this research and the lessons from COVID-19 create opportunities to advance considerations on the incorporation of the full effects of vaccines and other health-protecting and health-promoting interventions in HTAs.

## Data Availability

Aggregate data generated or analysed during this study are available from the corresponding author, upon review.

## Supplementary Materials

The supplementary material includes Appendix A, Appendix B, Appendix C, Appendix D and Appendix E.

## Author Contributions

Conceptualisation, M.D.F, K.M., D.M., J.R., G.S.; methodology, M.D.F, K.M., D.M., J.R., G.S.; software, M.D.F, D.M., K.M., J.R., G.S.; validation, L.S., D.E.B., M.D., K.H., F.R.J., J.P.S., R.P., D.S., A.T.; formal analysis, M.D.F, D.M., K.M., J.Y, G.S.; investigation, M.D.F, D.M., J.R., J.Y., K.M.; resources, M.D.F; data curation, K.M., M.D.F, D.M., J.Y., G.S.; writing—original draft preparation, M.D.F., K.M., G.S., and Shailja Vaghela; writing—review and editing, M.D.F., D.M., L.S., D.E.B., M.D., K.H., F.R.J., J.P.S., R.P., D.S., A.T., J.R., G.S., J.Y., K.M..; visualisation, M.D.F., D.M., J.Y., K.M., G.S.; supervision, M.D.F; project administration, M.D.F., D.M., G.S., J.Y., K.M.; funding acquisition, M.D.F. All authors have read and agreed to the published version of the manuscript.

## Institutional Review Board Statement

Not applicable.

## Informed Consent Statement

Not applicable.

## Acknowledgments

The authors acknowledge Professor F. Reed Johnson (Population Health Science, Duke School of Medicine) and Professor Flavio Toxvaerd (Faculty of Economics, University of Cambridge) for their participation in the expert panels. The authors also acknowledge Elizabeth Hamson, Mary M Moran, Carole Czudek (Pfizer employees) and Gyorgyi Feldmajer (Evidera employees) for specific contributions to this research project. Editorial support was provided by Shailja Vaghela of HealthEcon Consulting, Inc and was funded by Pfizer.

## Conflicts of Interest

MDF, DM, JY are employees of Pfizer and may hold stock or stock options of Pfizer. KM, JR and GS are employees of Evidera, which received financial support from Pfizer, Inc. in connection with the study and the development of this manuscript. Shailja Vaghela is an employee of HealthEcon Consulting, Inc. and an external consultant for Pfizer who has received consulting fees from Pfizer in connection with the development of this manuscript.

L.S. is an employee of the Office of Health Economics, a registered charity and Independent Research Organisation which receives funding from a variety of private and public sector sources.

D.E.B. consults for Data for Decisions, LLC on research contracts with Pfizer. He has also received compensation from Pfizer, Sanofi, GSK, and Merck for providing consulting services and for speaking and participating in meetings, executive education programs, and advisory boards.

M.D. has consulted on several economic evaluations of vaccines for different manufacturers

K.H. received personal fees from GSK and WHO for research unrelated to this work

J.P.S. is partner and head of Health Analytics at Lane Clark & Peacock LLP, Chair of the Royal Society for Public Health and reports personal fees from Novo Nordisk A/S and Pfizer Ltd outside of the submitted work.

D.S. Paid consultancies for Pfizer, GSK, Sanofi, AstraZeneca, Clover, Moderna, MSD, BioNTech, Seqirus.

A.T. is an employee of the Office of Health Economics, a registered charity and Independent Research Organisation which receives funding from a variety of private and public sector sources.

The experts were compensated for their time during the expert discussions.

## References

1. Mofijur M, Fattah IMR, Alam MA, et al. Impact of COVID-19 on the social, economic, environmental and energy domains: Lessons learnt from a global pandemic. Sustainable Production and Consumption. 2021;26:343–59.

2. Nicola M, Alsafi Z, Sohrabi C, et al. The socio-economic implications of the coronavirus pandemic (COVID-19): A review. Int J Surg. 2020;78:185–93.

3. Richards F, Kodjamanova P, Chen X, et al. Economic Burden of COVID-19: A Systematic Review. Clinicoecon Outcomes Res. 2022;14:293–307.

4. Kaye AD, Okeagu CN, Pham AD, et al. Economic impact of COVID-19 pandemic on healthcare facilities and systems: International perspectives. Best Pract Res Clin Anaesthesiol. 2021;35(3):293–306.

5. Saladino V, Algeri D, Auriemma V. The Psychological and Social Impact of Covid-19: New Perspectives of Well-Being. Frontiers in Psychology. 2020;11.

6. Hartley DM, Perencevich EN. Public Health Interventions for COVID-19: Emerging Evidence and Implications for an Evolving Public Health Crisis. JAMA. 2020;323(19):1908–9.

7. Ayouni I, Maatoug J, Dhouib W, et al. Effective public health measures to mitigate the spread of COVID-19: a systematic review. BMC Public Health. 2021;21(1):1015.

8. Neville FG, Templeton A, Smith JR, Louis WR. Social norms, social identities and the COVID-19 pandemic: Theory and recommendations. Social and Personality Psychology Compass. 2021;15(5):e12596.

9. Liu S, Zhu J, Liu Y, et al. Perception of strong social norms during the COVID-19 pandemic is linked to positive psychological outcomes. BMC Public Health. 2022;22(1):1403.

10. Advisory Committee on Immunization Practice (ACIP). ACIP Evidence to Recommendation User’s Guide 2020 [Available from: https://www.cdc.gov/vaccines/acip/recs/grade/downloads/acip-evidence-rec-frame-user-guide.pdf.

11. Joint Committee on Vaccination and Immunisation (JCVI). Code of Practice June 2013 2013 [Available from: https://assets.publishing.service.gov.uk/government/uploads/system/uploads/attachment_data/file/224864/JCVI_Code_of_Practice_revision_2013_-_final.pdf.

12. National Institute for Health and Care Excellence (NICE). Guide to the methods of technology appraisal 2013. NICE; 2013.

13. Bell E, Neri M, Steuten L. Towards a Broader Assessment of Value in Vaccines: The BRAVE Way Forward. Appl Health Econ Health Policy. 2022;20(1):105–17.

14. Asukai Y, Briggs A, Garrison LP, et al. Principles of Economic Evaluation in a Pandemic Setting: An Expert Panel Discussion on Value Assessment During the Coronavirus Disease 2019 Pandemic. Pharmacoeconomics. 2021;39(11):1201–8.

15. Barnighausen T, Bloom DE, Cafiero-Fonseca ET, O’Brien JC. Valuing vaccination. Proc Natl Acad Sci U S A. 2014;111(34):12313–9.

16. Bloom DE, Brenzel L, Cadarette D, Sullivan J. Moving beyond traditional valuation of vaccination: Needs and opportunities. Vaccine. 2017;35 Suppl 1:A29–a35.

17. Brassel S, Neri, M., and Steuten, L.,. Realising the Value of Vaccines in the UK: Ready for Prime Time? OHE Consulting Report, London: Office of Health Economics 2021 [Available from: https://www.ohe.org/publications/realising-broader-value-vaccines-uk-ready-prime-time.

18. Brassel S, Neri M, Schirrmacher H, Steuten L. The Value of Vaccines in Maintaining Health System Capacity in England. Office of Health Economics; 2021.

19. Brassel S, Steuten L. The Broader Value of Vaccines: The Return on Investment From a Governmental Perspective. Office of Health Economics; 2020.

20. Deogaonkar R, Hutubessy R, van der Putten I, Evers S, Jit M. Systematic review of studies evaluating the broader economic impact of vaccination in low and middle income countries. BMC Public Health. 2012;12:878.

21. Department of Health and Social Care (DHSC). Cost-effectiveness methodology for vaccination programmes. Consultation on the Cost-Effectiveness Methodology for Vaccination Programmes and Procurement (CEMIPP) Report 2018 [Available from: https://assets.publishing.service.gov.uk/government/uploads/system/uploads/attachment_data/file/707847/cemipp-consultation-document.pdf.

22. Goring S GL, Jansen JP, Briggs AH. Navigating the publishing landscape: novel elements of the value flower: fake or truly novel? Value Outcomes Spotlight. 2021;7(3).

23. Jit M, Hutubessy R. Methodological Challenges to Economic Evaluations of Vaccines: Is a Common Approach Still Possible? Appl Health Econ Health Policy. 2016;14(3):245–52.

24. Lakdawalla DN, Doshi JA, Garrison LP, Jr., et al. Defining Elements of Value in Health Care-A Health Economics Approach: An ISPOR Special Task Force Report [3]. Value Health. 2018;21(2):131–9.

25. Mauskopf J, Standaert B, Connolly MP, et al. Economic Analysis of Vaccination Programs: An ISPOR Good Practices for Outcomes Research Task Force Report. Value Health. 2018;21(10):1133–49.

26. Postma M, Biundo E, Chicoye A, et al. Capturing the value of vaccination within health technology assessment and health economics: Country analysis and priority value concepts. Vaccine. 2022;40(30):3999–4007.

27. Sanders GD, Neumann PJ, Basu A, et al. Recommendations for Conduct, Methodological Practices, and Reporting of Cost-effectiveness Analyses: Second Panel on Cost-Effectiveness in Health and Medicine. JAMA. 2016;316(10):1093–103.

28. Bloom DE, Cadarette D, Ferranna M. The Societal Value of Vaccination in the Age of COVID-19. Am J Public Health. 2021;111(6):1049–54.

29. Standaert B, Sauboin C, DeAntonio R, et al. How to assess for the full economic value of vaccines? From past to present, drawing lessons for the future. J Mark Access Health Policy. 2020;8(1):1719588.

30. Rob Johnson BD, David Haw, Patrick Doohan, Giovanni Forchin, Matteo Pianella, Neil Ferguson, Peter C Smith, and Katharina D Hauck. Report 51 - Valuing lives, education and the economy in an epidemic: Societal benefit of SARS-CoV-2 booster vaccinations in Indonesia. 2022.

31. McMillan SS, King M, Tully MP. How to use the nominal group and Delphi techniques. Int J Clin Pharm. 2016;38(3):655–62.

32. Nair R, Aggarwal R, Khanna D. Methods of formal consensus in classification/diagnostic criteria and guideline development. Semin Arthritis Rheum. 2011;41(2):95–105.

33. Taylor E. We Agree, Don’t We? The Delphi Method for Health Environments Research. HERD. 2020;13(1):11–23.

34. Grisham T. The Delphi technique: a method for testing complex and multifaceted topics. International Journal of Managing Projects in Business. 2009;2(1):112–30.

35. Mullen PM. Delphi: myths and reality. Journal of Health Organization and Management. 2003;17(1):37–52.

36. Fitch K, Bernstein SJ, Aguilar MD, et al. The RAND/UCLA Appropriateness Method User’s Manual. Santa Monica, CA: RAND Corporation; 2001.

37. Diamond IR, Grant RC, Feldman BM, et al. Defining consensus: a systematic review recommends methodologic criteria for reporting of Delphi studies. J Clin Epidemiol. 2014;67(4):401–9.

38. Henderson EJ, Rubin GP. Development of a community-based model for respiratory care services. BMC Health Serv Res. 2012;12:193.

39. Slade SC, Dionne CE, Underwood M, Buchbinder R. Standardised method for reporting exercise programmes: protocol for a modified Delphi study. BMJ Open. 2014;4(12):e006682.

40. Cutler DM, Summers LH. The COVID-19 Pandemic and the $16 Trillion Virus. JAMA. 2020;324(15):1495–6.

41. Department of Health and Social Care (DHSC), Office for National Statistics (ONS). Direct and Indirect Health Impacts of COVID-19 in England: Gov.UK; September 2021 [Available from: https://www.gov.uk/government/publications/dhsc-direct-and-indirect-health-impacts-of-covid-19-in-england-long-paper-9-september-2021.

42. Kirson N, Swallow E, Lu J, et al. The societal economic value of COVID-19 vaccines in the United States. J Med Econ. 2022;25(1):119–28.

43. Mayo M, Potugari B, Bzeih R, et al. Cancer Screening During the COVID-19 Pandemic: A Systematic Review and Meta-analysis. Mayo Clin Proc Innov Qual Outcomes. 2021;5(6):1109–17.

44. Buehrle DJ, Nguyen MH, Wagener MM, Clancy CJ. Impact of the Coronavirus Disease 2019 Pandemic on Outpatient Antibiotic Prescriptions in the United States. Open Forum Infectious Diseases. 2020;7(12).

45. Langford BJ, So M, Raybardhan S, et al. Antibiotic prescribing in patients with COVID-19: rapid review and meta-analysis. Clin Microbiol Infect. 2021;27(4):520–31.

46. Lane Clark & Peacock (LCP). Hidden health needs ‘the elephant in the NHS waiting room’ as waiting list number could rise to over 15 million in 2023 2021 [Available from: https://www.lcp.uk.com/media-centre/2021/12/hidden-health-needs-the-elephant-in-the-nhs-waiting-room-as-waiting-list-number-could-rise-to-over-15-million-in-2023/.

47. Pecetta S, Tortorice D, Scorza FB, et al. The trillion dollar vaccine gap. Sci Transl Med. 2022;14(638):eabn4342.

48. Bloom DE, Fan VY, Sevilla JP. The broad socioeconomic benefits of vaccination. Sci Transl Med. 2018;10(441).

49. Ferranna MaR, Lisa A and Cadarette, Daniel and Eber, Michael and Bloom David E. The Benefits and Costs of U.S. Employer COVID-19 Vaccine Mandates. National Bureau of Economic Research; 2022.

50. Congressional Budget Office. Budgetary Effects of the 2020 Coronavirus Pandemic 2020 [Available from: https://www.cbo.gov/publication/56388.

51. Heald D, Hodges R. The accounting, budgeting and fiscal impact of COVID-19 on the United Kingdom. Journal of Public Budgeting, Accounting & Financial Management. 2020.

52. Arnon A, Ricco J, Smetters K. Epidemiological and economic effects of lockdown. Brookings Papers on Economic Activity. 2020;2020(3):61–108.

53. Chen J, Vullikanti A, Santos J, et al. Epidemiological and Economic Impact of COVID-19 in the US. Scientific reports. 2021;11(1):1–12.

54. Choi Y, Kim H-j, Lee Y. Economic consequences of the COVID-19 pandemic: will it be a barrier to achieving sustainability? Sustainability. 2022;14(3):1629.

55. Chudik A, Mohaddes K, Pesaran MH, Raissi M, Rebucci A. A counterfactual economic analysis of Covid-19 using a threshold augmented multi-country model. J Int Money Finance. 2021;119:102477.

56. Hanushek EA, Woessmann L. The economic impacts of learning losses. 2020.

57. Keogh-Brown MR, Jensen HT, Edmunds WJ, Smith RD. The impact of Covid-19, associated behaviours and policies on the UK economy: A computable general equilibrium model. SSM Popul Health. 2020;12:100651.

58. Penn Wharton, University of Pennsylvania. COVID-19 Learning Loss: Long-run Macroeconomic Effects Update 2021 [Available from: https://budgetmodel.wharton.upenn.edu/issues/2021/10/27/covid-19-learning-loss-long-run-macro-effects.

59. Sandmann FG, Davies NG, Vassall A, et al. The potential health and economic value of SARS-CoV-2 vaccination alongside physical distancing in the UK: a transmission model-based future scenario analysis and economic evaluation. Lancet Infect Dis. 2021;21(7):962–74.

60. Singh V, Mishra V. Environmental impacts of coronavirus disease 2019 (COVID-19). Bioresour Technol Rep. 2021;15:100744.

61. Foundation TH. Health and social care funding to 2024/25. Slide deck of key findings. 2021.

62. Nguyen M. The Psychological Benefits of COVID-19 Vaccination. Advances in Public Health. 2021;2021.

63. Venter ZS, Aunan K, Chowdhury S, Lelieveld J. COVID-19 lockdowns cause global air pollution declines. Proceedings of the National Academy of Sciences. 2020;117(32):18984–90.

64. Groves HE, Piché-Renaud P-P, Peci A, et al. The impact of the COVID-19 pandemic on influenza, respiratory syncytial virus, and other seasonal respiratory virus circulation in Canada: A population-based study. The Lancet Regional Health - Americas. 2021;1:100015.

65. Chow EJ, Uyeki TM, Chu HY. The effects of the COVID-19 pandemic on community respiratory virus activity. Nature Reviews Microbiology. 2022.

66. Huang QS, Wood T, Jelley L, et al. Impact of the COVID-19 nonpharmaceutical interventions on influenza and other respiratory viral infections in New Zealand. Nat Commun. 2021;12(1):1001.

67. van den Hout WB. The value of productivity: human-capital versus friction-cost method. Annals of the Rheumatic Diseases. 2010;69(Suppl 1):i89.

