## Supplementary material for "The Societal Value of Vaccines: Expert-Based Conceptual Framework and Methods Using COVID-19 Vaccines as A Case Study"

### APPENDIX A: Questionnaire for Round 1 of Individual Expert Elicitation

Thank you for taking the time to answer this survey.

This questionnaire aims to gather initial inputs and perspectives on the topics of the Expert Panel.

Please provide your responses after reviewing the pre-read materials.

Your insights will support preparations for the upcoming panel discussions. The responses will be anonymous and analysed at aggregated level; the results will be used solely for the purpose of this piece of research.

Thank you.

* Required

**Demographic questions**

1. In which country do you conduct research on? *
   - o UK
   - o US
   - o Other:

2. What is your functional expertise? (Can select multiple options)
   - o Health policy
   - o Epidemiology
   - o Health Technology Assessment
   - o Public Health
   - o Immunisation
   - o Public finance
   - o Patient advocacy group
   - o Health economics
   - o Paediatrics / Child health
   - o Other:

3. What type of organisation do you work in or advise?
   - Government Institution
   - Academic Institution e.g., University
   - Patient Association
   - Other:

4. For how many years have you worked in healthcare research or sector? *

5. Have you applied a broader perspective in your prior work? *

NB. This question applies only to experts with previous experience of economic evaluations (e.g., health economists), please select 'Not applicable' otherwise.

- No
- Yes, in prior vaccine evaluations only
- Yes, in vaccine and non-vaccine specific evaluations
- Yes, in non-vaccine specific evaluations only
- Not applicable
- Other

**Effects of COVID-19 on broader economic and societal outcomes**

**Research objective: to identify the key societal outcomes impacted by COVID-19 and value elements**

6. Please rank the value elements below according to their relevance or priority for inclusion in economic evaluations of vaccines against COVID- 19 (1 low, 5 high) (1/2) *

Definitions of the elements of value of vaccination below are provided in the pre-read part 1 (slides 19-22).

Please state your opinion independently of the perspective recommended by HTA bodies in your country.

| | 1 | 2 | 3 | 4 | 5 |
|---|---|---|---|---|---|
| A1. Impact on length of life and quality of life of patients | ○ | ○ | ○ | ○ | ○ |
| B1.1 Impact on quality of life of carers | ○ | ○ | ○ | ○ | ○ |
| B1.2 Impact on quality of life of other individuals | ○ | ○ | ○ | ○ | ○ |
| B2. Transmission | ○ | ○ | ○ | ○ | ○ |
| B3. Burden of disease | ○ | ○ | ○ | ○ | ○ |
| B4. Value to other interventions | ○ | ○ | ○ | ○ | ○ |
| B5. AMR Prevention | ○ | ○ | ○ | ○ | ○ |
| B6. Mental health impact | ○ | ○ | ○ | ○ | ○ |
| B7. Health impact of congestion externality | ○ | ○ | ○ | ○ | ○ |

| | 1 | 2 | 3 | 4 | 5 |
|---|---|---|---|---|---|
| B8. Health equity | ◯ | ◯ | ◯ | ◯ | ◯ |
| C1.1. Avoided care cost of infected patients | ◯ | ◯ | ◯ | ◯ | ◯ |
| C1.2. Avoided care costs related to broad health effects | ◯ | ◯ | ◯ | ◯ | ◯ |
| C2. Financial sustainability and programmatic synergies | ◯ | ◯ | ◯ | ◯ | ◯ |
| C3. Public sector budget impact | ◯ | ◯ | ◯ | ◯ | ◯ |
| D1.1 Impact on patient productivity | ◯ | ◯ | ◯ | ◯ | ◯ |
| D1.2 Impact on carer productivity | ◯ | ◯ | ◯ | ◯ | ◯ |
| D1.3 Impact on productivity of other individuals | ◯ | ◯ | ◯ | ◯ | ◯ |
| D2. Impact on costs of non- pharmaceutic al interventions | ◯ | ◯ | ◯ | ◯ | ◯ |
| D3.1 Impact on foregone education of patient | ◯ | ◯ | ◯ | ◯ | ◯ |
| D3.2 Impact on foregone education of other individuals | ◯ | ◯ | ◯ | ◯ | ◯ |

7. Please rank the value elements below according to their relevance or priority for inclusion in economic evaluations of vaccines against COVID- 19 (1 low, 5 high) (2/2) *

Definitions of the elements of value of vaccination below are provided in the pre-read part 1 (slides 19-22).

Please state your opinion independently of the perspective recommended by HTA bodies in your country.

| | 1 | 2 | 3 | 4 | 5 |
|---|---|---|---|---|---|
| D4. Changes in household behaviour | ◯ | ◯ | ◯ | ◯ | ◯ |
| D5. Macroeconomic effects | ◯ | ◯ | ◯ | ◯ | ◯ |
| D6. Income equity | ◯ | ◯ | ◯ | ◯ | ◯ |
| D7. Scientific spill-over effects | ◯ | ◯ | ◯ | ◯ | ◯ |
| D8. Environmental Effects | ◯ | ◯ | ◯ | ◯ | ◯ |
| E1. Insurance Value | ◯ | ◯ | ◯ | ◯ | ◯ |
| E2. Real option value | ◯ | ◯ | ◯ | ◯ | ◯ |
| E3.1 Value of hope | ◯ | ◯ | ◯ | ◯ | ◯ |
| E3.2 Value of knowing | ◯ | ◯ | ◯ | ◯ | ◯ |
| E3.3 Fear of Diseases | ◯ | ◯ | ◯ | ◯ | ◯ |

8. What main criteria have you considered for prioritizing the elements of value above? *

9. In your opinion, are there any other elements of value not listed above which should be considered in economic evaluations of vaccines against COVID-19?

If so, please provide the rationale for inclusion and where you would rank them. *

Please state your opinion independently of the perspective recommended by HTA bodies in your country.

10. Please briefly explain if and why some of the elements of value above would NOT be conceptually appropriate to consider *

This question is only applicable where answer to previous questions is 'Yes, some outcomes above would not be appropriate '

11. In your opinion, what is the quality of evidence* supporting the inclusion of outcomes listed below in vaccine assessments? (1/2) *

Please refer to the pre-read materials and rank each element of value from 1 (lowest) to 5 (highest).

*The assessment of a study quality includes contextual, pragmatic and methodological considerations to establish how near the 'truth' its findings are likely to be and if the findings are of relevance in the particular setting or patient group of interest. Quality assessment considers appropriateness of study design to the research objective, risk of bias, statistical issues, generalizability, among others (CRD, University of York, 2009) [1].

| | 1 | 2 | 3 | 4 | 5 |
|---|---|---|---|---|---|
| A1. Impact on length of life and quality of life of patients | ◯ | ◯ | ◯ | ◯ | ◯ |
| B1.1 Impact on quality of life of carers | ◯ | ◯ | ◯ | ◯ | ◯ |
| B1.2 Impact on quality of life of other individuals | ◯ | ◯ | ◯ | ◯ | ◯ |
| B2. Transmission | ◯ | ◯ | ◯ | ◯ | ◯ |
| B3. Burden of disease | ◯ | ◯ | ◯ | ◯ | ◯ |
| B4. Value to other interventions | ◯ | ◯ | ◯ | ◯ | ◯ |
| B5. AMR Prevention | ◯ | ◯ | ◯ | ◯ | ◯ |
| B6. Mental health impact | ◯ | ◯ | ◯ | ◯ | ◯ |
| B7. Health impact of congestion externality | ◯ | ◯ | ◯ | ◯ | ◯ |
| B8. Health equity | ◯ | ◯ | ◯ | ◯ | ◯ |
| C1.1. Avoided care cost of infected patients | ◯ | ◯ | ◯ | ◯ | ◯ |
| C1.2. Avoided care costs related to broad health effects | ◯ | ◯ | ◯ | ◯ | ◯ |
| C2. Financial sustainability and programmatic synergies | ◯ | ◯ | ◯ | ◯ | ◯ |
| C3. Public sector budget impact | ◯ | ◯ | ◯ | ◯ | ◯ |
| D1.1 Impact on patient productivity | ◯ | ◯ | ◯ | ◯ | ◯ |
| D1.2 Impact on carer productivity | ◯ | ◯ | ◯ | ◯ | ◯ |
| D1.3 Impact on productivity of other individuals | ◯ | ◯ | ◯ | ◯ | ◯ |
| D2. Impact on costs of non- pharmaceutic al interventions | ◯ | ◯ | ◯ | ◯ | ◯ |
| D3.1 Impact on foregone education of patient | ◯ | ◯ | ◯ | ◯ | ◯ |
| D3.2 Impact on foregone education of other individuals | ◯ | ◯ | ◯ | ◯ | ◯ |

12. In your opinion, what is the **quality of evidence*** supporting the inclusion of outcomes listed below in vaccine assessments? (2/2) *

Please refer to the pre-read materials and rank each element of value from 1 (lowest) to 5 (highest).

*The assessment of a study quality includes contextual, pragmatic and methodological considerations to establish how near the 'truth' its findings are likely to be and if the findings are of relevance in the particular setting or patient group of interest. Quality assessment considers appropriateness of study design to the research objective, risk of bias, statistical issues, generalizability, among others (CRD, University of York, 2009).

| | 1 | 2 | 3 | 4 | 5 |
|---|---|---|---|---|---|
| D4. Changes in household behaviour | ◯ | ◯ | ◯ | ◯ | ◯ |
| D5. Macroeconomic effects | ◯ | ◯ | ◯ | ◯ | ◯ |
| D6. Income equity | ◯ | ◯ | ◯ | ◯ | ◯ |
| D7. Scientific spill-over effects | ◯ | ◯ | ◯ | ◯ | ◯ |
| D8. Environmental Effects | ◯ | ◯ | ◯ | ◯ | ◯ |
| E1. Insurance Value | ◯ | ◯ | ◯ | ◯ | ◯ |
| E2. Real option value | ◯ | ◯ | ◯ | ◯ | ◯ |
| E3.1 Value of hope | ◯ | ◯ | ◯ | ◯ | ◯ |
| E3.2 Value of knowing | ◯ | ◯ | ◯ | ◯ | ◯ |
| E3.3 Fear of Diseases | ◯ | ◯ | ◯ | ◯ | ◯ |

13. In your opinion, is the inclusion of the outcomes listed below **likely feasible***? (1/2) *

Please refer to the pre-read materials and rank each element of value from 1 (lowest) to 5 (highest).

*Feasibility defined as: existing methodological approaches would allow the inclusion of societal outcomes in economic models

| | 1 | 2 | 3 | 4 | 5 |
|---|---|---|---|---|---|
| A1. Impact on length of life and quality of life of patients | ◯ | ◯ | ◯ | ◯ | ◯ |
| B1.1 Impact on quality of life of carers | ◯ | ◯ | ◯ | ◯ | ◯ |
| B1.2 Impact on quality of life of other individuals | ◯ | ◯ | ◯ | ◯ | ◯ |
| B2. Transmission | ◯ | ◯ | ◯ | ◯ | ◯ |
| B3. Burden of disease | ◯ | ◯ | ◯ | ◯ | ◯ |
| B4. Value to other interventions | ◯ | ◯ | ◯ | ◯ | ◯ |
| B5. AMR Prevention | ◯ | ◯ | ◯ | ◯ | ◯ |
| B6. Mental health impact | ◯ | ◯ | ◯ | ◯ | ◯ |
| B7. Health impact of congestion externality | ◯ | ◯ | ◯ | ◯ | ◯ |
| B8. Health equity | ◯ | ◯ | ◯ | ◯ | ◯ |
| C1.1. Avoided care cost of infected patients | ◯ | ◯ | ◯ | ◯ | ◯ |
| C1.2. Avoided care costs related to broad health effects | ◯ | ◯ | ◯ | ◯ | ◯ |
| C2. Financial sustainability and programmatic synergies | ◯ | ◯ | ◯ | ◯ | ◯ |
| C3. Public sector budget impact | ◯ | ◯ | ◯ | ◯ | ◯ |
| D1.1 Impact on patient productivity | ◯ | ◯ | ◯ | ◯ | ◯ |
| D1.2 Impact on carer productivity | ◯ | ◯ | ◯ | ◯ | ◯ |
| D1.3 Impact on productivity of other individuals | ◯ | ◯ | ◯ | ◯ | ◯ |

| | 1 | 2 | 3 | 4 | 5 |
|---|---|---|---|---|---|
| D2. Impact on costs of non- pharmaceutic al interventions | ◯ | ◯ | ◯ | ◯ | ◯ |
| D3.1 Impact on foregone education of patient | ◯ | ◯ | ◯ | ◯ | ◯ |
| D3.2 Impact on foregone education of other individuals | ◯ | ◯ | ◯ | ◯ | ◯ |

14. In your opinion, is the inclusion of the outcomes listed below **likely feasible***? (2/2) *

Please refer to the pre-read materials and rank each element of value from 1 (lowest) to 5 (highest).

*Feasibility defined as: existing methodological approaches would allow the inclusion of societal outcomes in economic models

| | 1 | 2 | 3 | 4 | 5 |
|---|---|---|---|---|---|
| D4. Changes in household behaviour | ◯ | ◯ | ◯ | ◯ | ◯ |
| D5. Macroeconomic effects | ◯ | ◯ | ◯ | ◯ | ◯ |
| D6. Income equity | ◯ | ◯ | ◯ | ◯ | ◯ |
| D7. Scientific spill-over effects | ◯ | ◯ | ◯ | ◯ | ◯ |
| D8. Environmental Effects | ◯ | ◯ | ◯ | ◯ | ◯ |
| E1. Insurance Value | ◯ | ◯ | ◯ | ◯ | ◯ |
| E2. Real option value | ◯ | ◯ | ◯ | ◯ | ◯ |
| E3.1 Value of hope | ◯ | ◯ | ◯ | ◯ | ◯ |
| E3.2 Value of knowing | ◯ | ◯ | ◯ | ◯ | ◯ |
| E3.3 Fear of Diseases | ◯ | ◯ | ◯ | ◯ | ◯ |

15. Which of the outcomes below are currently already routinely included within an assessment by HTA bodies in your country? (1/2)

For those not included, please select option for main rationale for exclusion *

Please state country of relevance in subsequent question.

| | Routinely Included in HTA | Not Included in HTA due to lack of robust supporting evidence | Not Included in HTA due to lack of data/difficult to quantify | Not Included in HTA due to lack of ability/capacity to assess | Not Included in HTA due to lack of willingness (not recognised as relevant or appropriate) | Don't know |
|---|---|---|---|---|---|---|
| A1. Impact on length of life and quality of life of patients | ◯ | ◯ | ◯ | ◯ | ◯ | ◯ |
| B1.1 Impact on quality of life of carers | ◯ | ◯ | ◯ | ◯ | ◯ | ◯ |
| B1.2 Impact on quality of life of other individuals | ◯ | ◯ | ◯ | ◯ | ◯ | ◯ |
| B2. Transmission | ◯ | ◯ | ◯ | ◯ | ◯ | ◯ |
| B3. Burden of disease | ◯ | ◯ | ◯ | ◯ | ◯ | ◯ |
| B4. Value to other interventions | ◯ | ◯ | ◯ | ◯ | ◯ | ◯ |
| B5. AMR Prevention | ◯ | ◯ | ◯ | ◯ | ◯ | ◯ |
| B6. Mental health impact | ◯ | ◯ | ◯ | ◯ | ◯ | ◯ |
| B7. Health impact of congestion externality | ◯ | ◯ | ◯ | ◯ | ◯ | ◯ |
| B8. Health equity | ◯ | ◯ | ◯ | ◯ | ◯ | ◯ |
| C1.1. Avoided care cost of infected patients | ◯ | ◯ | ◯ | ◯ | ◯ | ◯ |
| C1.2. Avoided care costs related to broad health effects | ◯ | ◯ | ◯ | ◯ | ◯ | ◯ |
| C2. Financial sustainability and programmatic synergies | ◯ | ◯ | ◯ | ◯ | ◯ | ◯ |
| C3. Public sector budget impact | ◯ | ◯ | ◯ | ◯ | ◯ | ◯ |
| D1.1 Impact on patient productivity | ◯ | ◯ | ◯ | ◯ | ◯ | ◯ |
| D1.2 Impact on carer productivity | ◯ | ◯ | ◯ | ◯ | ◯ | ◯ |
| D1.3 Impact on productivity of other individuals | ◯ | ◯ | ◯ | ◯ | ◯ | ◯ |
| D2. Impact on costs of non- pharmaceutic al interventions | ◯ | ◯ | ◯ | ◯ | ◯ | ◯ |
| D3.1 Impact on foregone education of patient | ◯ | ◯ | ◯ | ◯ | ◯ | ◯ |
| D3.2 Impact on foregone education of other individuals | ◯ | ◯ | ◯ | ◯ | ◯ | ◯ |

16. Which of the outcomes below are currently already routinely included within an assessment by HTA bodies in your country? (2/2)

For those not included, please select option for main rationale for exclusion *

Please state country of relevance in subsequent question

| | Routinely Included in HTA | Not Included in HTA due to lack of robust supporting evidence | Not Included in HTA due to lack of data/difficult to quantify | Not Included in HTA due to lack of ability/capacity to assess | Not Included in HTA due to lack of willingness (not recognised as relevant or appropriate) | Don't know |
|---|---|---|---|---|---|---|
| D4. Changes in household behaviour | ◯ | ◯ | ◯ | ◯ | ◯ | ◯ |
| D5. Macroeconomic effects | ◯ | ◯ | ◯ | ◯ | ◯ | ◯ |
| D6. Income equity | ◯ | ◯ | ◯ | ◯ | ◯ | ◯ |
| D7. Scientific spill-over effects | ◯ | ◯ | ◯ | ◯ | ◯ | ◯ |
| D8. Environmental Effects | ◯ | ◯ | ◯ | ◯ | ◯ | ◯ |
| E1. Insurance Value | ◯ | ◯ | ◯ | ◯ | ◯ | ◯ |
| E2. Real option value | ◯ | ◯ | ◯ | ◯ | ◯ | ◯ |
| E3.1 Value of hope | ◯ | ◯ | ◯ | ◯ | ◯ | ◯ |
| E3.2 Value of knowing | ◯ | ◯ | ◯ | ◯ | ◯ | ◯ |
| E3.3 Fear of Diseases | ◯ | ◯ | ◯ | ◯ | ◯ | ◯ |

17. Please enter country of reference for question 15/16. *

18. Are there any additional barriers to inclusion of the outcomes listed in previous question? If so, what could be potential solutions? *

19. Please share any examples of evidence or good practice in the broad value assessment of vaccines, relating to any of the outcomes above. *

These might be examples from your own country or anywhere else in the world. The examples will be used to help moderate the workshop discussions on overcoming barriers to the inclusion of key outcomes in value assessments of vaccines.

**Methods for inclusion of broader value elements**

**Research objective: to identify methodological approaches to include key societal outcomes impacted by COVID-19 in economic evaluations of vaccines**

20. Are the following suitable methods/approaches for inclusion of broader value elements within an assessment of the societal value of vaccination against COVID-19? *

Rank from 1 to 5 based on the relative appropriateness (1 low, 5 high). Methods listed below are described in pre-read part 1 (slides 29 to 31).

Please state your opinion independently of the perspective recommended by HTA bodies in your country.

| | 1 | 2 | 3 | 4 | 5 | Unsure |
|---|---|---|---|---|---|---|
| B5. Antimicrobial resistance (AMR)- extensions to CEA or CBA | ◯ | ◯ | ◯ | ◯ | ◯ | ◯ |
| B6. & C1.2. Mental health impact- Approach based on number of additional depression cases | ◯ | ◯ | ◯ | ◯ | ◯ | ◯ |
| B6. & C1.2. Mental health impact- Approach based on impact of vaccination on months spent in depression | ◯ | ◯ | ◯ | ◯ | ◯ | ◯ |
| B7. & C1.2 Congestion externality - Opportunity cost | ◯ | ◯ | ◯ | ◯ | ◯ | ◯ |
| C3. Public finance impact- return on investment (ROI) | ◯ | ◯ | ◯ | ◯ | ◯ | ◯ |
| C3. Public finance impact- fiscal benefit to cost ratio (fBCR) | ◯ | ◯ | ◯ | ◯ | ◯ | ◯ |
| D1. Productivity loss - Human capital | ◯ | ◯ | ◯ | ◯ | ◯ | ◯ |
| D1. Productivity loss - Friction cost | ◯ | ◯ | ◯ | ◯ | ◯ | ◯ |
| D2. Impact on the cost of non- pharmaceutic al interventions (NPIs)- Approach based on relationship between vaccination and NPI levels | ◯ | ◯ | ◯ | ◯ | ◯ | ◯ |
| D3. Education loss- Approach based on impact on test scores | ◯ | ◯ | ◯ | ◯ | ◯ | ◯ |
| D3. Education loss- Microsimulation | ◯ | ◯ | ◯ | ◯ | ◯ | ◯ |
| D5. Impact on GDP- Approach relies on external estimates for GDP | ◯ | ◯ | ◯ | ◯ | ◯ | ◯ |
| D5. Impact on GDP- Simple estimate using time series data | ◯ | ◯ | ◯ | ◯ | ◯ | ◯ |
| D5. Impact on GDP- Macroeconomic modelling | ◯ | ◯ | ◯ | ◯ | ◯ | ◯ |

21. Are there any methods not listed above that should be considered? *

Please state any relevant methods, respective value elements and envisaged advantages if applicable

22. If any method listed above would not be suitable or unclear, please explain why *

23. Open feedback on the Survey

Please share your feedback around the usability, design, wording, terminology and clarity of the questions in this questionnaire

### APPENDIX B: COVID-19 Impact Inventory

#### Table S1 Impact Inventory

| Original (Panel 1) CATEGORY | Report CATEGORY | Original (Panel 1) DEFINITION | Report definition | Change |
|---|---|---|---|---|
| **A. Narrow Health Effects** | **A. Narrow Health Effects** | Impact of vaccines on the health of vaccinated individuals | Impact of vaccines on the health of vaccinated individuals | - |
| **A1. Impact on length of life and QoL of patients** | **A1. Impact on length of life and QoL of patients** | Impact on life expectancy or life-years saved, and on patients' physical, mental, emotional, and social functioning | Impact on life expectancy or life-years saved, and on patients' physical, mental, emotional, and social functioning, including mortality and QoL impact of potential adverse events related to vaccination. Definition based on Deogaonkar (2012) | Definition expanded to include QoL impact of adverse events related to vaccination |
| **B. Broad Health Effects** | **B. Broad Health Effects** | Impact of vaccines on the health of the unvaccinated population | Impact of vaccines on the health of the unvaccinated population | - |
| **B1. Impact on QoL**<br>**B1.1 Impact on QoL of carers**<br>**B1.2 Impact on QoL of other individuals** | **B1. Impact on QoL**<br>**B1.1 Impact on QoL of carers**<br>**B1.2 Impact on QoL of other individuals** | Impact on caregivers' and other individuals' physical, mental, emotional, and social functioning | Impact on caregivers' and other individuals' physical, mental, emotional, and social functioning | - |
| **B2. Transmission value** | **B2. Transmission value** | Impact on disease transmission patterns and associated morbidity and mortality | Impact on disease transmission patterns and associated morbidity and mortality | - |
| **B3. Burden of disease** | **B3. Burden of disease** | Impact on overall burden of disease to society, in terms of prevalence and severity, estimated through the total amount of associated morbidity and mortality<br>(Note, this includes A1., B1., and B2.) | Impact on overall burden of disease to society, in terms of prevalence and severity, estimated through the total amount of associated morbidity and mortality<br>(Note, this includes A1., B1., and B2.) | - |
| **B4. Value to other interventions** | **B4. Value to other interventions** | Impact on the cost effectiveness of other non-vaccine interventions (also referred to as enablement value) | Impact on the cost effectiveness of other non-vaccine interventions (also referred to as enablement value) | -<br>Note, that in manuscript it is called: enablement value / value to other interventions |

| Original (Panel 1) CATEGORY | Report CATEGORY | Original (Panel 1) DEFINITION | Report definition | Change |
|---|---|---|---|---|
| **B5. AMR prevention value** | **B5. AMR prevention value** | Impact on the rate of development and transmission of resistant bacterial infections, and associated morbidity and mortality | Impact on the rate of development and transmission of resistant bacterial infections, and associated morbidity and mortality | - |
| **B6. Mental health impact** | **B6. Mental health impact** | Impact on mental health and well-being of the population through avoiding non-pharmaceutical interventions impacting mental health (e.g., lockdowns, school closures) | Impact on mental health and well-being of the population directly through vaccination and indirectly through avoiding non-pharmaceutical interventions that can impact mental health (e.g., lockdowns, school closures) | Definition changed to include direct effect |
| **B7. Health impact of congestion externality** | **B7. Health system impact** | Impact on morbidity and mortality in the population through avoiding or mitigating overload of public health service facilities and resulting delays in diagnostic and care services | Impact of vaccination on morbidity and mortality in the population through avoiding or mitigating overload of public health service facilities and resulting delays in diagnostic and care services | Category name changed, definition slightly changed |
| **B8. Health equity value** | **B8. Health equity value** | The absence of disparities in health | Vaccination's impact on disparities in health | Definition changed to emphasize vaccination |
| **C. Effect on public finances** | **C. Effect on public finances** | The costs of vaccination and its cost offsets to public finances | The costs of vaccination and its cost offsets to public finances | - |
| **C1. Cost offsets to health care system**<br>**C1.1 Avoided care cost of infected patients**<br>**C1.2 Avoided care cost related to broad health effects** | **C1. Cost offsets to health care system**<br>**C1.1 Avoided care cost of infected patients**<br>**C1.2 Avoided care cost related to broad health effects** | As with many interventions, vaccines come at a cost to the health care system and also generate cost offsets by avoiding downstream health care consumption.<br>• C1.1 The value of avoiding the excess costs of treatment of more severe cases<br>• C1.2 The value of avoiding costs related to broad health effects including mental health care costs and extra care costs related to delayed diagnosis and care due to congestion externality | As with many interventions, vaccines come at a cost to the health care system and also generate cost offsets by avoiding downstream health care consumption.<br>• C1.1 The value of avoiding the excess costs of treatment of more severe cases<br>• C1.2 The value of avoiding costs related to broad health effects including mental health care costs and extra care costs related to delayed diagnosis and care due to health system impact | - |
| **C2. Financial sustainability and programmatic synergies** | **C2. Financial sustainability and programmatic synergies** | Improved financial sustainability of health care programs as a result of synergies with vaccination programs and/or stimulation of private demand. (Deogaonkar 2012) | Improved financial sustainability of health care programs as a result of synergies with vaccination programs and/or stimulation of private demand. (Definition based on Deogaonkar, 2012) | - |
| **C3. Public sector budget impact** | **C3. Public sector budget impact** | Impact on government revenues (e.g., taxes and social security contributions) and expenditures (e.g., transfers including sick benefit) related to | Impact on government revenues (e.g., taxes and social security contributions) and expenditures (e.g., transfers including sick benefit) related to | - |

| Original (Panel 1) CATEGORY | Report CATEGORY | Original (Panel 1) DEFINITION | Report definition | Change |
|---|---|---|---|---|
| | | the productivity impact (D1) and macroeconomic effects (D5) corresponding to the effect of vaccination on morbidity and mortality, and on the level of non-pharmaceutical interventions. | the productivity impact (D1) and macroeconomic effects (D5) corresponding to the effect of vaccination on morbidity and mortality, and on the level of non-pharmaceutical interventions. | |
| **D. Societal and economic effects** | **D. Societal and economic effects** | Economic impact of vaccines outside of the public sector | Economic impact of vaccines outside of the public sector | - |
| **D1. Productivity impact**<br>**D1.1 Impact on patient productivity**<br>**D1.2 Impact on carer productivity**<br>**D1.3 Impact on productivity of other individuals** | **D1. Productivity impact**<br>**D1.1 Impact on patient productivity**<br>**D1.2 Impact on carer productivity**<br>**D1.3 Impact on productivity of other individuals** | • D1.1 Impact on lost days of work and on the level of productivity at work, both for getting vaccinated and for disease or mortality avoided<br>• D1.2 Impact on caregivers' time spent and level of productivity at work due to caring for a patient or taking them to be vaccinated<br>• D1.3 Impact on lost days of work and reduced productivity through contribution to avoiding non-pharmaceutical interventions (e.g., lockdowns preventing work or impacting work efficiency, school closures decreasing parents' working hours and productivity) | • D1.1 Impact on lost days of work and on the level of productivity at work, both for getting vaccinated and for disease or mortality avoided<br>• D1.2 Impact on caregivers' time spent and level of productivity at work due to caring for a patient or taking them to be vaccinated<br>• D1.3 Impact on lost days of work and reduced productivity through contribution to avoiding non-pharmaceutical interventions (e.g., lockdowns preventing work or impacting work efficiency, school closures decreasing parents' working hours and productivity) | - |
| **D2. Impact on costs of non-pharmaceutical interventions** | **D2. Impact on costs of non-pharmaceutical interventions** | Reduction or elimination of the need for, and hence the costs of non-pharmaceutical interventions designed to contain disease outbreaks, epidemics, or pandemics (e.g., lockdowns, use of face masks) | Reduction or elimination of the need for, and hence the costs of non-pharmaceutical interventions designed to contain disease outbreaks, epidemics, or pandemics (e.g., lockdowns, use of face masks) | - |
| **D3. Impact on foregone education**<br>**D3.1 Impact on foregone education of patient**<br>**D3.2 Impact on foregone education of other individuals** | **D3. Impact on foregone education**<br>**D3.1 Impact on foregone education of patient**<br>**D3.2 Impact on foregone education of other individuals** | Contribution to the avoidance of lost school days due to illness or school closures related to disease containment measures | Contribution to the avoidance of lost school days directly due to illness or indirectly through school closures (related to disease containment measures) | Definition changed to include direct / indirect effects |

| Original (Panel 1) CATEGORY | Report CATEGORY | Original (Panel 1) DEFINITION | Report definition | Change |
|---|---|---|---|---|
| **D4. Changes in household behaviour** | **D4. Changes in individual and household behaviour** | Economic improvements due to changes in household choices such as fertility and consumption/savings as a result of vaccination | Consequences of changes in household choices as a result of vaccination, including areas of fertility, consumption/savings, attitude towards risk of infection and infection control measures, willingness to vaccinate against other diseases | Both category name and definition changed to reflect individual behaviour as well |
| **D5. Macroeconomic effects** | **D5. Macroeconomic effects** | Reduction or elimination of the macroeconomic impact of lost productivity and non-pharmaceutical interventions designed to contain disease outbreaks, epidemics, or pandemics (Note, macroeconomic effects are affected by D4.) | Reduction or elimination of the macroeconomic impact of lost productivity and non-pharmaceutical interventions designed to contain disease outbreaks, epidemics, or pandemics (Note, macroeconomic effects are affected by D4.) | - |
| **D6. Income equity value** | **D6. Income equity value** | Reduction or elimination of the impact of the disease and of non-pharmaceutical interventions designed to contain it on the income distribution | Reduction or elimination of the impact of the disease and of non-pharmaceutical interventions designed to contain it on the income distribution | - |
| **D7. Scientific spill-over effects** | **D7. Scientific spill-over effects** | The impact of research and development on our collective knowledge, arising when innovators cannot entirely appropriate the benefit of scientific advances | The impact of research and development on our collective knowledge, arising when innovators (vaccine developers) cannot entirely appropriate the benefit of scientific advances | Minor definition change |
| **D8. Environmental effects** | **D8. Environmental effects** | The effect the additional waste generated by vaccination exerts on the environment, and the effect on air and water pollution, and waste generation through impact on productivity and the level of non-pharmaceutical interventions, including widespread use of disposable items | The effect the additional waste generated by vaccination exerts on the environment, and the effect on air and water pollution, and waste generation through impact on productivity and the level of non-pharmaceutical interventions, including widespread use of disposable items | - |
| **E. Uncertainty value** | **E. Uncertainty value** | The values generated by different concepts revolving around uncertainty. | The values generated by different concepts revolving around uncertainty. | - |
| **E1. Insurance value** | **E1. Insurance value** | The value to vaccinated individuals of being protected from the physical and financial burden of an illness. It has two components: vaccination reduces the 'physical risk' of getting sick and vaccination expands the possibilities for insuring against illness ('financial risk protection') - Lakdawalla (2018) | The value to vaccinated individuals of being protected from the physical and financial burden of an illness. It has two components: vaccination reduces the 'physical risk' of getting sick and vaccination expands the possibilities for insuring against illness ('financial risk protection') - Definition based on Lakdawalla et al 2018 | - |
| **E2. Real option value** | **E2. Real option value** | Opportunities created for the patient to benefit from future advances in medicine by extending their life | Opportunities created for the patient to benefit from future advances in medicine by extending their life Definition based on Lakdawalla et al 2018 | - |

| Original (Panel 1) CATEGORY | Report CATEGORY | Original (Panel 1) DEFINITION | Report definition | Change |
|---|---|---|---|---|
| **E3. Psychological benefits related to reduced uncertainty**<br>**E3.1 Value of hope**<br>**E3.2 Value of knowing**<br>**E3.3 Fear of diseases / contagion** | **E3. Psychological effects related to uncertainty**<br>**E3.1 Value of hope**<br>**E3.2 Value of knowing**<br>**E3.3 Fear of diseases / contagion**<br>**E3.4 Vaccine anxiety** | • E3.1 Impact of reducing probability of illness on patients' utility who may value a treatment/intervention with high variability in outcomes (e.g., a severely ill patient undertaking a risky procedure for a low probability chance of a cure) or may prefer a treatment/intervention with less variability around expected outcomes (e.g., a COVID-19 vaccine lowering the chance of hospitalization)<br>• E3.2 Impact on patients' utility who may attach value from the knowledge that a certain diagnosis will predict treatment effectiveness.<br>• E3.3 The value of reducing the anxiety of a (future) spread of a disease | • E3.1 Impact of reducing probability of illness on patients' utility who may value an intervention with high variability in outcomes (e.g., a severely ill patient undertaking a risky procedure for a low probability chance of a cure) or may prefer a treatment/intervention with less variability around expected outcomes (e.g., a COVID-19 vaccine lowering the chance of hospitalization) Definition based on Lakdawalla et al 2018<br>• E3.2 Impact on patients' utility who may attach value to the knowledge that a vaccine will impact the probability of illness and severe outcomes Definition based on Lakdawalla et al 2018<br>• E3.3 The value of reducing the anxiety of a (future) spread of a disease Definition based on Lakdawalla et al 2018<br>• E3.4 The negative psychological effect related to the actual and/or perceived risk of vaccine adverse effects | E3.4 Vaccine anxiety added, E3.2 definition changed for clarity |

AMR, antimicrobial resistance; COVID-19, coronavirus disease 2019; DALY, disability-adjusted life years; GDP, gross domestic product; QALY, quality-adjusted life years; QoL, quality of life; R&D, research and development

### APPENDIX C: 'Quality of Evidence' linking the value elements to COVID-19 and/or vaccination and 'Ability to include' the value elements in the framework

The evaluation was based on a targeted literature review and subjective judgment, as opposed to systematic literature reviews, and strictly defined standardised criteria. It should only be regarded as an indicative starting point for expert discussion. Similar evaluations were presented for certain value elements in Jit *et al*. [2], and the OHE Consulting Report 2021 [3], where expert opinion was inferred using the Delphi method.

As shown in the Table S2, Ability reflected the availability of methods for inclusion in quantitative vaccine assessments – even value elements without monetisable value can be reflected in qualitative assessments. The evidence identified was in some cases empirical, whereas in other cases only based on model estimates. While all evidence are subject to uncertainties arising from potential measurement errors, estimates from economic models are based on underlying model assumptions, which is an additional source of uncertainty. Further, literature is quickly evolving, hence the low level of evidence for certain value items may improve over time.

**Table S2 Evidence Appraisal**

| | | | | |
|---|---|---|---|---|
| **Evidence:** is there good quality evidence about causal pathways to broader impact? | • Likelihood of causal relationship based on available identified evidence and/or expert judgment<br>• Quality of identified empirical evidence<br>• Quantitative size of the effect | **High (1)**<br>Strong causal relationship, high quality of evidence for COVID impact AND vaccine effect | **Moderate (2)**<br>Moderate causal relationship and moderate quality of evidence for COVID OR vaccine effect | **Low (3)**<br>Very limited evidence and unclear direction, small or no effects identified |
| **Ability:** is it feasible to estimate the $/QALY impact? | • Availability of monetary value of the element<br>• Timeframe over which the effect is expected to manifest (short-term usually has lower uncertainty) | **High (1)**<br>Can be monetised (method exists) | **Moderate (2)**<br>Can be monetised (method exists) but with strong assumptions and/or long term only | **Low (3)**<br>No monetary value or very high uncertainty |

COVID-19, coronavirus disease 2019; QALY, quality-adjusted life years

### APPENDIX D: Summary of the evidence review and gap analysis

#### Figure S1 Evidence Review and Gap Analysis (1/3)

| BRAVE | VALUE CATEGORY | DESCRIPTION | BRAVE (BELL 2022) | EVIDENCE | ABILITY |
|---|---|---|---|---|---|
| A | **A. Narrow Health Effects** | **Impact of vaccines on the health of vaccinated individuals** | | | |
| | **A1. Impact on length of life and QoL of patients** | Impact of vaccines on life expectancy or life-years saved, and on patients' physical, mental, emotional, and social functioning | √ | 1 | 1 |
| B | **B. Broad Health Effects** | **Impact of vaccines on the health of the unvaccinated population** | | | |
| | **B1. Impact on QoL**<br>**B1.1 Impact on QoL of carers**<br>**B1.2 Impact on QoL of other individuals** | Impact of vaccines on caregivers' and other individuals' physical, mental, emotional, and social functioning | √<br>N | 1<br>3 | 1<br>2 |
| | **B2. Transmission value** | Impact of vaccination on disease transmission patterns and associated morbidity and mortality | √ | 1 | 1 |
| | **B3. Burden of disease** | Impact on overall burden of disease to society, in terms of prevalence and severity, estimated through the total amount of associated morbidity and mortality (Note, this includes A1., B1., and B2.) | √ | 1 | 1 |
| | **B4. Value to other interventions / enablement value** | Impact of vaccination on the cost effectiveness of other non-vaccine interventions | √ | 3 | 2 |
| | **B5. AMR prevention value** | Impact on the rate of development and transmission of resistant bacterial infections, and associated morbidity and mortality. Preventing infectious disease through vaccination reduces antibiotic use and therefore reduces antimicrobial resistance. | √ | 3 | 3 |
| | **B6. Mental health impact** | Impact of vaccination on mental health and well-being of the population through avoiding non-pharmaceutical interventions impacting mental health (e.g., lockdowns, school closures). This may in turn also reduce prevalence of other disorders (anxiety, substance abuse, stress disorders) that are exacerbated by a pandemic. | Not explicitly | 1 | 1 |
| | **B7. Health system impact** | Impact on morbidity and mortality in the population through avoiding or mitigating overload of public health service facilities and resulting delays in diagnostic and care services | N | 1 | 2 |
| | **B8. Health equity value** | Vaccination can lead to more equal distribution of health outcomes. | N | 2 | 3 |

High (1) | Moderate (2) | Low (3)

AMR, antimicrobial resistance; QoL, quality of life

### Figure S2 Evidence Review and Gap analysis (2/3)

| BRAVE | VALUE CATEGORY | DESCRIPTION | BRAVE (BELL 2022) | EVIDENCE | ABILITY |
|---|---|---|---|---|---|
| C | **C. Effect on public finances** | **The costs of vaccination and its cost offsets to public finances** | | | |
| | **C1. Cost offsets to health care system**<br>**C1.1 Avoided care cost of infected patients**<br>**C1.2 Avoided care cost related to broad health effects** | C1. Impact on medical costs borne by the health system from potential reductions in the number of general practitioner and specialist consultations, treatment, screening interventions, and hospitalizations<br>C1.1 The value of avoiding the excess costs of treatment of more severe cases<br>C1.2 The value of avoiding costs related to broad health effects including mental health care costs and extra care costs related to delayed diagnosis and care due to congestion externality | √<br>N | 1<br>2 | 1<br>2 |
| | **C2. Financial sustainability and programmatic synergies** | Improved financial sustainability of health care programs as a result of synergies with vaccination programs and/or stimulation of private demand. | N | 3 | 1 |
| | **C3. Public sector budget impact** | Impact on government revenues (e.g., taxes and social security contributions) and expenditures (e.g., transfers including sick benefit) related to the productivity impact (D1) and macroeconomic effects (D5) corresponding to the effect of vaccination on morbidity and mortality, and on the level of non-pharmaceutical interventions. | √ | 1 | 2 |
| D | **D. Societal and economic effects** | **Economic impact of vaccines outside of the public sector** | | | |
| | **D1. Productivity impact**<br>**D1.1 Impact on patient productivity**<br>**D1.2 Impact on carer productivity**<br>**D1.3 Impact on productivity of other individuals** | D1.1 Impact on lost days of work and on the level of productivity at work, both for getting vaccinated and for disease or mortality avoided<br>D1.2 Impact on caregivers' time spent and level of productivity at work due to caring for a patient or taking them to be vaccinated<br>D1.3 Impact on lost days of work and reduced productivity through contribution to avoiding non-pharmaceutical interventions (e.g., lockdowns preventing work or impacting work efficiency, school closures decreasing parents' working hours and productivity) | √<br>√<br>N | 1<br>1<br>2 | 1<br>1<br>2 |
| | **D2. Impact on costs of non-pharmaceutical interventions** | Reduction or elimination of the need for, and hence the costs of non-pharmaceutical interventions designed to contain disease outbreaks, epidemics, or pandemics (e.g., lockdowns, use of face masks) | N | 3 | 2 |
| | **D3. Impact on foregone education**<br>**D3.1 Impact on foregone education of patient**<br>**D3.2 Impact on foregone education of other individuals** | Contribution to the avoidance of lost school days due to illness or school closures related to disease containment measures | N<br>N | 1<br>2 | 3<br>3 |

High (1) | Moderate (2) | Low (3)

### Figure S3 Evidence Review and Gap Analysis (3/3)

| BRAVE | VALUE CATEGORY | DESCRIPTION | BRAVE (BELL 2022) | EVIDENCE | ABILITY |
|---|---|---|---|---|---|
| D | **D4. Changes in household behaviour** | Economic improvements due to changes in household choices such as fertility and consumption/savings as a result of vaccination | N | 3 | 3 |
| | **D5. Macroeconomic effects** | Reduction or elimination of the macroeconomic impact of lost productivity and non-pharmaceutical interventions designed to contain disease outbreaks, epidemics, or pandemics (Note, macroeconomic effects are affected by D4.) | √ | 1 | 2 |
| | **D6. Income equity value** | Reduction or elimination of the impact of the disease and of non-pharmaceutical interventions designed to contain it on the income distribution | √ | 2 | 3 |
| | **D7. Scientific spill-over effects** | The impact of research and development on our collective knowledge, arising when innovators cannot entirely appropriate the benefit of scientific advances | N | 3 | 3 |
| | **D8. Environmental effects** | The effect the additional waste generated by vaccination exerts on the environment, and the effect on air and water pollution, and waste generation through impact on productivity and the level of non-pharmaceutical interventions, including widespread use of disposable items | N | 2 | 2 |
| E | **E. Uncertainty value** | The values generated by different concepts revolving around uncertainty. | | | |
| | **E1. Insurance value** | The value to vaccinated individuals of being protected from the physical and financial burden of an illness. It has two components: vaccination reduces the 'physical risk' of getting sick and vaccination expands the possibilities for insuring against illness ('financial risk protection') | N | 3 | 2 |
| | **E2. Real option value** | Vaccination's impact on opportunities created for the patient to benefit from future advances in medicine by extending their life | N | 3 | 3 |
| NEW | **E3. Psychological benefits related to reduced uncertainty**<br>**E3.1 Value of hope**<br>**E3.2 Value of knowing**<br>**E3.3 Fear of diseases / contagion** | E3.1 Impact of reducing probability of illness on patients' utility who may value a treatment/intervention with high variability in outcomes (e.g., a severely ill patient undertaking a risky procedure for a low probability chance of a cure) or may prefer a treatment/intervention with less variability around expected outcomes (e.g., a COVID-19 vaccine lowering the chance of hospitalization)<br>E3.2 Impact on patients' utility who may attach value to the knowledge that a certain diagnosis will predict treatment effectiveness.<br>E3.3 The value of reducing the anxiety of a (future) spread of a disease | N<br>N<br>Not explicitly | 3<br>3<br>3 | 3<br>3<br>3 |

High (1) | Moderate (2) | Low (3)

COVID-19, coronavirus disease 2019

### APPENDIX E: Identified Quantification Methods for Broader Value Elements and Expert Elicitation on their Appropriateness

**Table S3 Quantification methods and Expert Elicitation**

<table>
<tr><th rowspan="2">Broader Value Element and Quantification Method - Literature Findings</th><th colspan="2">Round 1 Polling (N=8)</th><th rowspan="2">Experts’ Views during the Panel 2 Discussions</th></tr>
<tr><th>% High rank (4 or 5)</th><th>% Low rank (1 or 2)</th></tr>
<tr><td colspan="4">B: Broader Health Effects; C: Effects on Public Finances</td></tr>
<tr><td>B5. AMR – extensions to CEA or CBA<br>Although the pandemic has influenced antibiotic use, both directly through antibiotics rx or indirectly by influencing access to antibiotics of other patients [4], no method was identified that has been applied to the quantification of the relationship between the pandemic and AMR.</td><td>50%</td><td>25%</td><td rowspan="3">- Experts stated that although antimicrobial resistance (AMR) was not ranked high, it might emerge in the future debates as new evidence will be available.</td></tr>
<tr><td>B6. & C1.2. Mental health impact – Approach based on number of additional depression cases<br>The mental health impact of the pandemic and the related social restrictions were widely documented, two approaches were identified, wherein, monetary value was attached to the mental impact by multiplying excess case numbers by average cost of treatment per person [5].</td><td>38%</td><td>50%</td></tr>
<tr><td>B6. & C1.2. Mental health impact – Approach based on impact of vaccination on months spent in depression<br>In the second approach, authors multiplied the similarly calculated severity-specific excess depression case numbers by QoL, and healthcare cost impact associated with each depression severity category [6].</td><td>38%</td><td>50%</td></tr>
<tr><td>B7. & C1.2 Health system impact – Opportunity cost</td><td>50%</td><td>50%</td><td></td></tr>
</table>

| Broader Value Element and Quantification Method - Literature Findings | Round 1 Polling (N=8) | | Experts' Views during the Panel 2 Discussions |
|---|---|---|---|
| | % High rank (4 or 5) | % Low rank (1 or 2) | |
| The disruption in service provision has been documented via a variety of outcome measures including backlogs, waiting times, the share of incomplete patient pathways etc. [3, 7-9] or on the demand side such as an estimate on the number of people that need care but have not yet come forward to receive care due to the pandemic [7]. Monetising the impact of this congestion externality used the foregone net monetary benefit associated with the treatments as the opportunity cost associated with inpatient bed-days [10]. | | | - The methodology for assessing the monetary value of the health system impact by evaluating healthcare resources by their opportunity cost as opposed to their accounting costs was considered appropriate and could be extended further to cover foregone screenings. This direct approach concentrated on the effect of the congestion externality and covered both mortality and QoL implications.<br>- Using excess deaths as a measure for the indirect mortality impact of COVID-19—including but not limited to the effect of health system congestion—is controversial. It is a simple but powerful method for capturing indirect mortality impact in a comprehensive way, and during periods where no other major change occurred that could substantially impact mortality, it is reasonable to assign all change in mortality to the pandemic. Some of the major confounding factors involved in these analyses, such as the reaping effect and mortality displacement may completely or partially be adjusted for by age standardisation and other statistical techniques.<br>- In forward looking analyses, a multiplier capturing the relationship between ICU case numbers and excess inpatient deaths could be used for predicting excess deaths based on case numbers.<br>- Excess deaths should only be assumed during time periods when demand for health services exceeds capacity. |
| While certain elements of the public finance impact of the pandemic have been estimated for both the US [11] and UK [12], no study was identified calculating the full public finance impact or the return on investment of the COVID-19 vaccination. | | | |
| **C3. Public finance impact – ROI** | 63% | 13% | |
| **C3. Public finance impact – fBCR** | 50% | 13% | |
| ***D: Societal and Economic Effects*** | | | |
| **D1. Productivity loss - Human capital** | 50% | 13% | - Human capital approach captures lost income due to mortality and morbidity associated with a disease at an individual level, but at an economic level, the structure of the labour market needs to be accounted for. |
| **D1. Productivity loss - Friction cost** | 50% | 25% | |

| Broader Value Element and Quantification Method - Literature Findings | Round 1 Polling (N=8) | | Experts' Views during the Panel 2 Discussions |
|---|---|---|---|
| | % High rank (4 or 5) | % Low rank (1 or 2) | |
| **D2. Impact on the cost of NPIs – Approach based on relationship between vaccination and NPI levels**<br><br>No identified work estimated the impact of vaccination on the direct cost of NPIs. A possible approach to quantifying this impact is to assess the change in resource use (e.g., in the number of face masks used) as a result of vaccination and multiply that by the unit cost. | 38% | 25% | - The friction cost method usually gives a smaller value estimate and is more appropriate in most situations.<br>- The loss of firm-specific human capital associated with losing job and getting re-employed is potentially bigger than the loss associated with time spent off work and hence needs to be accounted for, alongside cross-country differences in labour market conditions.<br>- Aspects of productivity are not generally considered in assessments include the reorganisation of production resulting from the pandemic, and the fact that losing one's job and getting re-employed is associated with a substantial loss of firm-specific human capital. |
| **D3. Education loss – Approach based on impact on test scores**<br><br>The OECD approach for assessing the impact of lost education on individual income and GDP [13], based on the impact of school closures on test scores (proxy for lost cognitive ability), and its relation to decrease in lifetime earnings. | 63% | 38% | - OECD approach was considered straightforward and worthy of conduction.<br>- Distributional consequences were also considered important to capture, and impacts need to be assessed across different education levels too.<br>-In assessments including macroeconomic impact, the estimated GDP impact associated with school closures needs to be reconciled with GDP changes arising from other factors to avoid double counting.<br>- Besides school closures, online education periods may also be accounted for by proxying the reduction in efficacy of online schooling compared to offline schooling based on assessments of work from home capabilities based on telecommunication infrastructure. |
| **D3. Education loss – Microsimulation**<br><br>The Penn Wharton Budget Model, an individual-level stochastic simulation model of labour productivity, in which the effect of lost education is incorporated through grade-specific achievement score gains that are converted into an estimate of loss in effective years of schooling [14]. | 75% | 13% | |
| **D5. Impact on GDP – Approach relies on external estimates for GDP** | 38% | 13% | - Avoiding NPIs and concentrating on GDP (or gross value added) as the only macroeconomic outcome measure is an appropriate |

| Broader Value Element and Quantification Method - Literature Findings | Round 1 Polling (N=8) | | Experts' Views during the Panel 2 Discussions |
|---|---|---|---|
| | % High rank (4 or 5) | % Low rank (1 or 2) | |
| Literature assessing the various aspects of the COVID-19 pandemic's macroeconomic impact was widely documented, mainly concentrating on GDP and employment as outcome measures, with 3 approaches. Certain studies relied on publicly available GDP estimates from other entities, such as the Congressional Budget Office [11] or Goldman Sachs [6]. | | | approach for synthesising macroeconomic impact without double-counting. It can be done either by directly estimating a multiplier of case numbers on GDP directly or by taking a two-step approach.<br>- The two-step approach involves estimating the multiplier between inpatient case numbers and NPI levels to capture the policy responses to decreasing case numbers as the policy is expected to be more responsive to impact on hospital capacities than on mortality, and then estimating another multiplier between NPI levels and GVA.<br>- When assessing impact on production and value added, it is important to cover non-market production too.<br>- Besides overall GDP, that is not sensitive to distributional outcomes, measures of macroeconomic performance should also consider impact on health equity and income inequalities through social welfare measures. While macroeconomic impact is important under pandemic settings, building de novo macroeconomic or combined epidemiological and macroeconomic models do not seem appropriate for HTA purposes, due to their complexity and inherent uncertainties. |
| **D5. Impact on GDP – Simple estimate using time series data** | 38% | 25% | |
| Other works performed simple time-series analyses comparing pre- and post-pandemic GDP levels [15]. | | | |
| **D5. Impact on GDP – Macroeconomic modelling** | 63% | 0% | |
| Based on the macroeconomic models, such as computable general equilibrium models or combined epidemiological and macroeconomic models [16-18]. | | | |
| **D8. Environmental effects** | NR | NR | |
| Various aspects of the environmental impact of the pandemic have been documented in the literature [19]. Positive impacts, such as air quality improvement, reduction in water and noise pollution, or getting closer to sustainable development goals are arising from the lower industrial activity and less traffic related to the disruption in economic activities. Negative impacts include biomedical waste generation, increased municipal waste generation, and a | | | |

| Broader Value Element and Quantification Method - Literature Findings | Round 1 Polling (N=8) | | Experts' Views during the Panel 2 Discussions |
|---|---|---|---|
| | % High rank (4 or 5) | % Low rank (1 or 2) | |
| parallel decrease in waste recycling in certain countries. No identified study estimated the monetary impact of these elements. | | | |
| ***E: Uncertainty Value*** | | | |
| **E3. Psychological effects** | NR | NR | - Psychological effects of vaccinations encompassing both the positive and negative elements, are important to account for in epidemiology models as they have an important impact on uptake and hence on infection numbers.<br>- Vaccine mandates impose cost on people through vaccine anxiety that should be included at least qualitatively but potentially even quantitatively. Including their direct QALY impact would increase complexity, likely without substantially changing conclusions.<br>- In some US states payments were offered for taking the vaccine, but these policies did not achieve big impact, suggesting the anxiety to potentially be substantive - which can be a topic for further research.<br>- Methods for quantifying insurance value and AMR prevention value are under development but no established best practice is available. For measuring vaccine anxiety, both stated preference and revealed preference approaches may be feasible. |
| Increasing rates of the COVID-19 vaccination rates have shown psychological benefits, measured by lower levels of anxiety, worry, displeasure, and depression in the US. However, no identified study had attached monetary value to the positive or negative psychological value of vaccination. | | | |

AMR, antimicrobial resistance; CBA, cost benefit analysis; CEA, cost effectiveness analysis; COVID-19, coronavirus disease 2019; DALY, disability-adjusted life years; fBCR, fiscal benefit to cost ratio; GDP, gross domestic product; GVA, gross value added; QALY, quality-adjusted life years; QoL, quality of life; NPI, non-pharmaceutical intervention; NR, not reported; OECD, Organisation for Economic Co-operation and Development; ROI, return on investment; UK, United Kingdom; US, United States

### REFERENCES

1. Centre for Reviews and Dissemination (CRD), Systematic reviews: CRD's guidance for undertaking reviews in health care. 2009.
2. Jit M, Hutubessy R. Methodological Challenges to Economic Evaluations of Vaccines: Is a Common Approach Still Possible? Appl Health Econ Health Policy. 2016;14(3):245-52.
3. Brassel S, Neri, M., and Steuten, L.,. Realising the Value of Vaccines in the UK: Ready for Prime Time? OHE Consulting Report, London: Office of Health Economics 2021 [Available from: https://www.ohe.org/publications/realising-broader-value-vaccines-uk-ready-prime-time .
4. Sevilla JP, Bloom DE, Cadarette D, Jit M, Lipsitch M. Toward economic evaluation of the value of vaccines and other health technologies in addressing AMR. Proceedings of the National Academy of Sciences. 2018;115(51):12911-9.
5. Cutler DM, Summers LH. The COVID-19 Pandemic and the $16 Trillion Virus. JAMA. 2020;324(15):1495-6.
6. Kirson N, Swallow E, Lu J, et al. The societal economic value of COVID-19 vaccines in the United States. J Med Econ. 2022;25(1):119-28.
7. Lane Clark & Peacock (LCP). Hidden health needs 'the elephant in the NHS waiting room' as waiting list number could rise to over 15 million in 2023 2021 [Available from: https://www.lcp.uk.com/media-centre/2021/12/hidden-health-needs-the-elephant-in-the-nhs-waiting-room-as-waiting-list-number-could-rise-to-over-15-million-in-2023/.
8. Mayo M, Potugari B, Bzeih R, et al. Cancer Screening During the COVID-19 Pandemic: A Systematic Review and Meta-analysis. Mayo Clin Proc Innov Qual Outcomes. 2021;5(6):1109-17.
9. The Health Foundation. Health and social care funding to 2024/25. Slide deck of key findings 2021 [Available from: https://www.health.org.uk/publications/reports/health-and-social-care-funding-to-2024-25.
10. Brassel S, Neri M, Schirrmacher H, Steuten L. The Value of Vaccines in Maintaining Health System Capacity in England. Office of Health Economics; 2021.
11. Congressional Budget Office. Budgetary Effects of the 2020 Coronavirus Pandemic 2020 [Available from: https://www.cbo.gov/publication/56388.
12. Heald D, Hodges R. The accounting, budgeting and fiscal impact of COVID-19 on the United Kingdom. Journal of Public Budgeting, Accounting & Financial Management. 2020.
13. Hanushek EA, Woessmann L. The economic impacts of learning losses. 2020.
14. Penn Wharton, University of Pennsylvania. COVID-19 Learning Loss: Long-run Macroeconomic Effects Update 2021 [Available from: https://budgetmodel.wharton.upenn.edu/issues/2021/10/27/covid-19-learning-loss-long-run-macro-effects.
15. Sandmann FG, Davies NG, Vassall A, et al. The potential health and economic value of SARS-CoV-2 vaccination alongside physical distancing in the UK: a transmission model-based future scenario analysis and economic evaluation. Lancet Infect Dis. 2021;21(7):962-74.
16. Arnon A, Ricco J, Smetters K. Epidemiological and economic effects of lockdown. Brookings Papers on Economic Activity. 2020;2020(3):61-108.
17. Choi Y, Kim H-j, Lee Y. Economic consequences of the COVID-19 pandemic: will it be a barrier to achieving sustainability? Sustainability. 2022;14(3):1629.
18. Chudik A, Mohaddes K, Pesaran MH, Raissi M, Rebucci A. A counterfactual economic analysis of Covid-19 using a threshold augmented multi-country model. J Int Money Finance. 2021;119:102477.
19. Singh V, Mishra V. Environmental impacts of coronavirus disease 2019 (COVID-19). Bioresour Technol Rep. 2021;15:100744.
